# Within-scan MRI signal and texture in Alzheimer’s disease: a multicohort study

**DOI:** 10.64898/2026.11.20.26368593

**Authors:** Wontack Han, Hyun-Jee Sarah Han, Dong-Joo Kim, Byung C. Yoon, the Alzheimer’s Disease Neuroimaging Initiative

**Author notes:** Corresponding author: Byung Chul Yoon, MD, PhD, E-mail address, VA Palo Alto Healthcare System, 3801 Miranda Ave. Palo Alto, CA 94304, USA. Declarations of interest: None. Data used in preparation of this article were obtained from the Alzheimer’s Disease Neuroimaging Initiative (ADNI) database (adni.loni.usc.edu). As such, the investigators within the ADNI contributed to the design and implementation of ADNI and/or provided data but did not participate in the analysis or writing of this report. A complete listing of ADNI investigators can be found at: https://adni.loni.usc.edu/wp-content/uploads/how_to_apply/ADNI_Acknowledgement_List.pdf.

## Abstract

**INTRODUCTION:** Conventional MRI contains tissue signal and texture information beyond regional volume, but acquisition variability limits their comparability. We evaluated within-scan signal and texture analyses for estimating contemporaneous cognition and amyloid/tau burden in Alzheimer’s disease (AD) beyond volumetry.

**METHODS:** We retrospectively analyzed 2,437 baseline datasets from ADNI3, ADNI4, MCSA, SCAN/CLARiTI, and OASIS-3. SynthSeg defined regions for T1-weighted and fluid-attenuated inversion recovery (FLAIR) measurements. Within-scan regional-to-rest log ratios (WSRLR) were compared with scanner-matched, cross-fitted cognitively normal (CN) references. Texture analysis used three-dimensional gray-level co-occurrence matrices (GLCM). Covariate-adjusted rank models examined associations; ADNI3-trained ridge, elastic-net, gradient-boosting, and TabPFN-v2 models were tested externally.

**RESULTS:** Moderate-to-strong correlations were observed between regional signal changes assessed using WSRLR vs. CN-reference (cohort median regional ρ=0.504–0.890; n=2,314). Lower entorhinal FLAIR WSRLR and greater entropy were associated with AD dementia adjusted for volume, while opposite correlations were observed with the hippocampus (all q<0.001). Greater entorhinal FLAIR entropy correlated with lower Montreal Cognitive Assessment (MoCA) and Mini-Mental State Examination (MMSE) scores (r=−0.198 and −0.098, respectively; both q<0.001). Lower entorhinal FLAIR WSRLR and T1 cluster shade were associated with higher regional tau uptake beyond volume and diagnosis (r=−0.146 and −0.205; both q≤0.010). Adding entorhinal and hippocampal signal and texture significantly improved pooled external MoCA (n=868) and MMSE (n=551) prediction in three of four models.

**DISCUSSION:** Within-scan signal and texture demonstrated AD-related regional associations and improved prediction of contemporaneous cognitive performance beyond volumetry. Prospective validation is needed to evaluate their potential value in opportunistic dementia screening.

## 1. Introduction

Cognitive impairment can be challenging to recognize in clinical practice, potentially delaying evaluation of Alzheimer’s disease (AD) and related dementias.[1,2] Conventional brain MRI plays a central role in AD evaluation by identifying alternative causes of cognitive impairment, assessing vascular contributions, and characterizing regional atrophy. However, atrophy patterns may not be disease-specific, and their absence does not exclude underlying AD pathology, limiting identification based on structural changes alone.[3] Beyond regional volume, the same images contain regional intensity differences and spatial patterns, or texture, that may provide additional tissue information. For instance, regional fluid-attenuated inversion recovery (FLAIR) and T2 signal abnormalities were associated with alterations in myelin and axonal density, with myelin content and cerebrovascular pathology contributing to white-matter signal variation.[4] Quantitative T1 mapping with tissue measurements also demonstrated contributions of myelin and iron to MRI contrast.[5] Quantifying these features could complement volumetry and support investigation of opportunistic cognitive assessment.

Building on these MRI-histopathologic correlations, studies have examined associations between MRI signal and texture in AD. In a Dominantly inherited Alzheimer Network (DIAN) study, Rahmani et al. reported associations between standardized T1/FLAIR intensity distributions and tau positron emission tomography (PET), including lower mean FLAIR signal and greater signal variability after accounting for regional atrophy.[6] Parietal white matter hyperintensity burden was also shown to predict dementia, predominantly attributed to AD, after accounting for hippocampal volume.[7] For textural analysis, a previous study on 150 Alzheimer Disease Neuroimaging Initiative (ADNI) participants showed correlations between regional T1-weighted gray-level co-occurrence (GLCM) matrix texture features and tau PET uptake.[8] Hippocampal texture differences were also found between cognitively unimpaired amyloid-positive and amyloid-negative individuals, and the incorporation of hippocampal texture provided incremental value for predicting cognitive change beyond hippocampal volume.[9]

Despite the potential value of MRI signal and texture analysis beyond volumetry, clinical application is complicated by acquisition parameters, receiver scaling, reconstruction, tissue composition, and partial-volume effects. Previous approaches used normative cohorts,[6] T1/T2 weighted ratios,[10] or anatomical references such as the callosal genu,[11] and cerebellar gray matter.[12] However, these strategies depend on comparable reference data, multiple image contrasts, or specific reference regions that may themselves be affected by disease. Herein, we evaluated the within-scan regional-to-rest log ratio (WSRLR) for relative characterization of regional T1-weighted and FLAIR signal. In WSRLR, the log-transformed median intensity of a target region for an individual sequence is contrasted with the median of the log-transformed regional median intensities from other brain regions. Drawing on log-ratio representations,[13] this approach characterizes relative T1 and FLAIR signal without normative data, cross-sequence measurements, or a single anatomical reference.

Complementary to regional signal assessments, GLCM analysis characterizes the spatial relationships between neighboring voxel intensities, providing texture features including entropy, contrast, autocorrelation, and cluster shade.[8] Entropy measures the uncertainty of the gray-level pair distribution, with higher values indicating a broader, less predictable mixture of voxel-pair combinations. Contrast measures the average gray-level differences between neighboring voxels. Autocorrelation measures the average product of neighboring gray levels, and cluster shade measures asymmetry in the distribution of paired gray-level sums around their mean.[8,14]

We combined WSRLR-based regional signal characterization with three-dimensional (3D) GLCM texture analysis of both T1-weighted and FLAIR images across five analytic groups: ADNI3,[15] ADNI4,[16] Mayo Clinic Study of Aging (MCSA),[17] Standardized Centralized Alzheimer’s and Related Dementias Neuroimaging (SCAN) and CLARiTI datasets accessed through the National Alzheimer’s Coordinating Center (NACC),[18,19] Open Access Series of Imaging Studies (OASIS-3).[20] We hypothesized that WSRLR enables relative regional signal characterization associated with regional volume, cognitive performance, amyloid/tau PET burden, and clinically attributed AD dementia, comparable to using reference data from cognitively normal (CN) populations. We further hypothesized that regional texture features would exhibit patterns of association with these AD-related outcomes that are distinct from those observed for relative signal intensity. Lastly, we hypothesized that signal and texture add incremental value to the prediction of contemporaneous cognitive performance beyond volumetry.

## 2. Methods

### 2.1. Study design and participants

This retrospective study used existing baseline MRI and linked clinical or PET data from five analytic groups: ADNI3, ADNI4, MCSA, SCAN/CLARiTI, and OASIS-3. NACC is the centralized data repository for the National Institute on Aging (NIA)-funded Alzheimer’s Disease Research Centers (ADRCs). SCAN is a multi-institutional initiative for standardized MRI and PET acquisition and analysis; NACC provides infrastructure for SCAN data integration and sharing. SCAN and CLARiTI imaging was linked to NACC clinical data and combined as one analytic group. The total number of participants included was 2,437, distributed across different analytic groups as follows: 658 ADNI3, 549 ADNI4, 747 Mayo, 408 SCAN/CLARiTI (325 SCAN and 83 CLARiTI), and 75 OASIS-3. ADNI data were accessed through the LONI Image and Data Archive (https://ida.loni.usc.edu/). Established in 2003 as a public–private partnership under the leadership of Michael W. Weiner, MD, ADNI integrates longitudinal imaging, biomarker, and clinical assessments to investigate AD progression. ADNI3 MRI data were downloaded on September 10 and 15, 2026, and linked clinical and PET tables were obtained from the August 5, 2026 export. ADNI4 imaging and linked clinical and PET tables were downloaded on September 23, 2026. One eligible baseline MRI was processed from each participant. ADNI4 records from 187 participants already represented in ADNI3 were excluded to prevent overlap between the training and test samples. ADNI4 participants with anti-amyloid therapy recorded in the available ADNI treatment table were excluded. Inclusion criteria included the availability of paired 3D T1 and FLAIR imaging. Participants were classified as CN, having mild cognitive impairment (MCI), or having dementia, using available cohort diagnoses. The AD-vs-CN comparison included only dementia clinically attributed to AD and CN participants; MCI and other dementias were not combined with the AD group. The overview of the study design is illustrated in Figure 1. This study was performed only with publicly available datasets, and local Institutional Review Board approval was not required.

**Figure 1.**
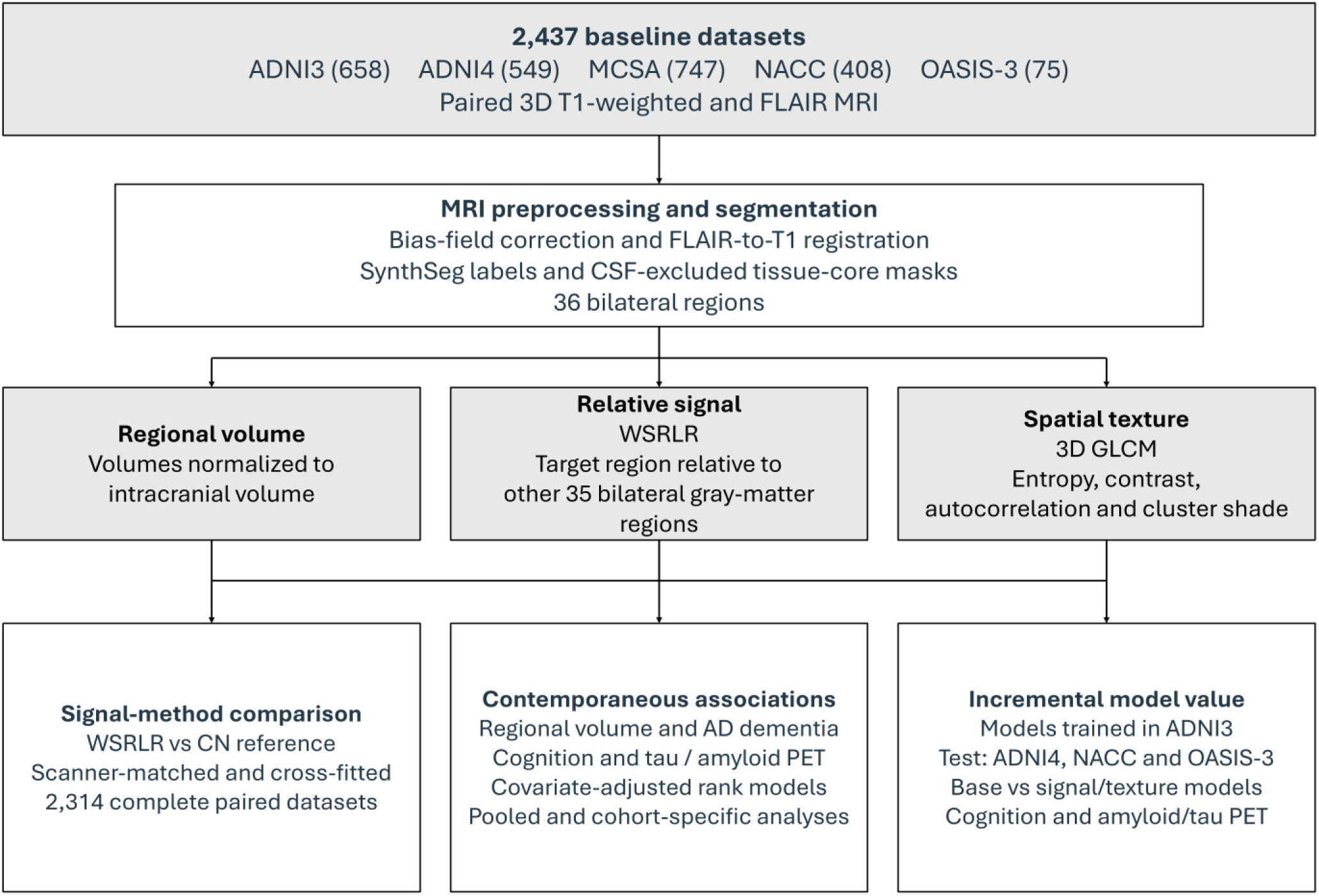
Retrospective, multi-cohort study design. WSRLR referenced each target to the other 35 of 36 bilateral gray-matter regions. Abbreviations: 3D, three-dimensional; AD, Alzheimer’s disease; ADNI3/4, Alzheimer’s Disease Neuroimaging Initiative, phases 3 and 4; CLARiTI, Consortium for Clarity in Alzheimer’s Disease and Related Dementias Research Through Imaging; CN, cognitively normal; CSF, cerebrospinal fluid; FLAIR, fluid-attenuated inversion recovery; GLCM, gray-level co-occurrence matrix; MCSA, Mayo Clinic Study of Aging; MRI, magnetic resonance imaging; NACC, National Alzheimer’s Coordinating Center; OASIS-3, Open Access Series of Imaging Studies, third release; PET, positron emission tomography; SCAN, Standardized Centralized Alzheimer’s and Related Dementias Neuroimaging; T1, longitudinal relaxation time; WSRLR, within-scan regional-to-rest log ratio.

### 2.2. MRI preprocessing and segmentation

MRIs were processed using 3D Slicer (version 5.12.3), including N4 bias-field correction,[21] and FLAIR images were registered to the corresponding T1-weighted images. Brain segmentation and cortical parcellation were performed using SynthSeg within FreeSurfer (version 8.0.0).[22] The analysis included 36 brain regions from each hemisphere, including 34 cortical parcels based on the Desikan-Killiany labeling scheme, as well as the hippocampus and amygdala (Supplementary Table 1). For primary analyses, homologous left and right regions were combined into 36 bilateral regions. Regional volumes were summed across hemispheres, and signal intensities were calculated as the median across voxels in the combined bilateral tissue-core masks. Total intracranial volume (ICV) was estimated automatically using SynthSeg[22] and was used to adjust for differences in head size. To reduce volume averaging with the cerebrospinal fluid (CSF), regional masks underwent one-voxel erosion using six-connected neighbors and exclusion of voxels within 1.5 mm of segmented CSF.

Quality control involved a combination of automated measurement-validity checks with visual review during pipeline development. Automated checks were used to assess image-grid consistency, finite positive intensities, retained tissue-core voxel counts of at least 200 voxels, and sufficient voxel pairs for texture extraction. A board-certified neuroradiologist (BCY) reviewed selected segmentation overlays and tissue-core masks for appropriate segmentation, particularly for cortical boundary errors and CSF inclusion. Regional measurements failing validity criteria were treated as missing.

### 2.3. Within-scan regional signal contrast

WSRLR was calculated by subtracting the median of the log-transformed regional median intensities across the reference panel from the log-transformed median intensity of the target region:

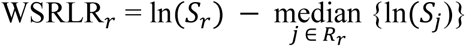

In this formula, S denotes the median intensity of the target region (“r”), and Rr denotes the reference regions. For each target, the reference comprised the other 35 bilateral gray-matter regions from the fixed 36-region panel (34 cortical parcels, hippocampus, and amygdala), with the target excluded from its own reference. A complete valid 36-region profile was required. These were calculated for each MRI sequence, T1 and FLAIR. A representative FLAIR slice and its corresponding regional WSRLR map are shown in Figure 2A. Whole-region measurements were mapped onto anatomical parcels for visualization.

**Figure 2.**
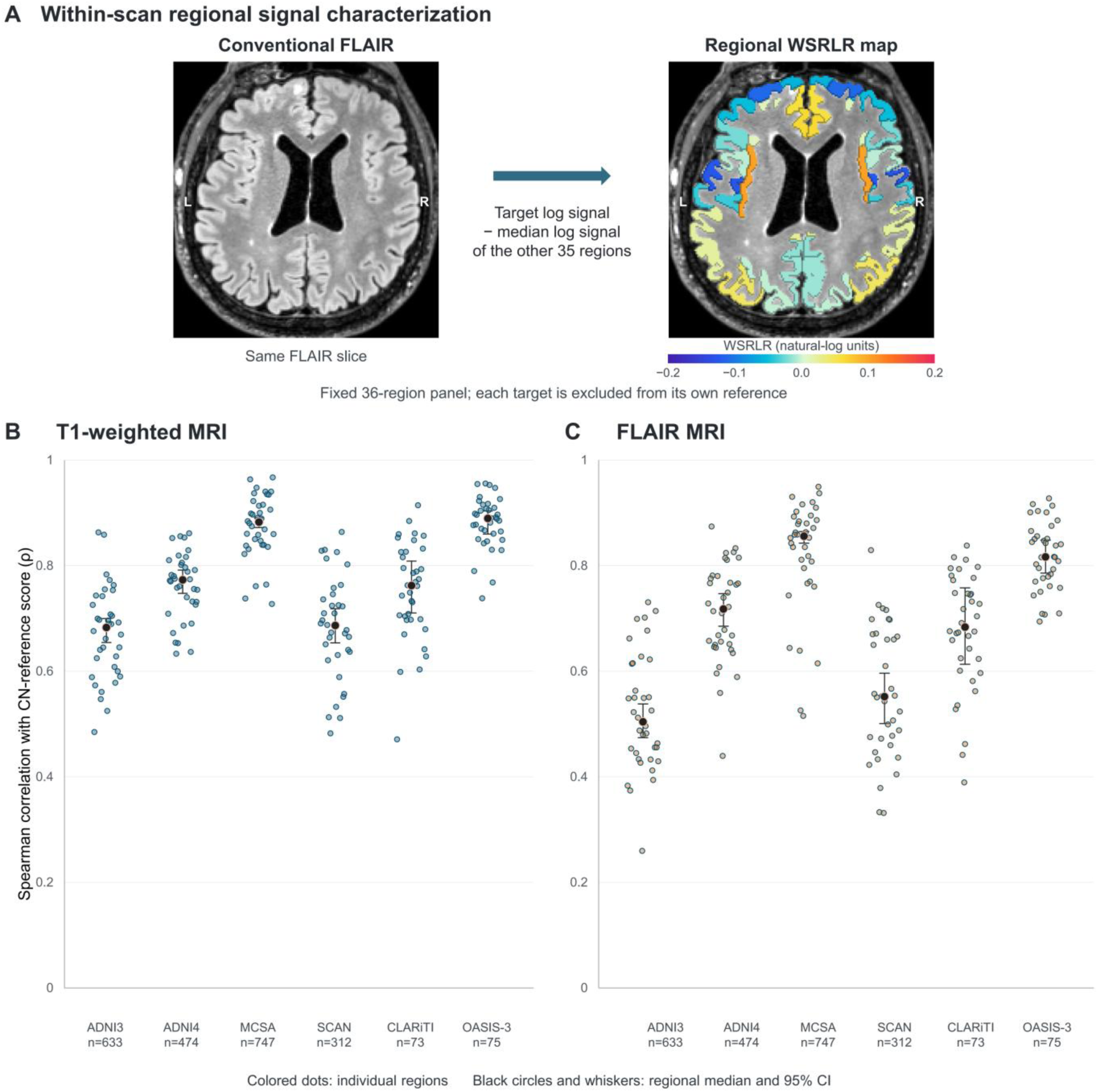
Within-scan regional signal characterization and correspondence with cognitively normal reference measurements. (A) A representative axial FLAIR slice and its corresponding regional WSRLR map. Each target region’s log-transformed median intensity was contrasted with the median log-transformed regional median intensities from the other 35 bilateral gray-matter regions. Measurements were obtained from three-dimensional, CSF-excluded tissue-core masks and mapped onto anatomical parcels for visualization. Colors reflect regional WSRLR values. (B, C) Within-cohort Spearman correlations between WSRLR and scanner-matched, cross-fitted CN-reference scores for T1-weighted and FLAIR MRI, respectively. Colored dots represent individual regions. Black circles indicate the median correlation across 36 regions; bars show participant-bootstrap 95% confidence intervals based on 1,000 resamples. Both sequences used 2,314 complete paired datasets. Abbreviations: CN, cognitively normal; CSF, cerebrospinal fluid; FLAIR, fluid-attenuated inversion recovery; MRI, magnetic resonance imaging; WSRLR, within-scan regional-to-rest log ratio.

### 2.4. Cognitively normal reference comparison

For comparison with WSRLR, CN-reference scores were derived from CN participants from the matching cohort, scanner manufacturer, model, and magnet strength. Regional intensities were first calibrated using cortical intensity distributions. CN data were then used to estimate expected regional intensities after accounting for age, sex, and available acquisition parameters. CN-reference scores reflected the differences between observed and expected intensities relative to reference variability. Participants were assigned to five groups, and CN-reference estimates for each group were derived from CN participants in the other four groups (five-fold cross-fitting). Scores were not calculated when scanner-matched CN data were insufficient. Comparisons between WSRLR and CN-reference scores were assessed using Spearman rank correlations.

### 2.5. Texture analysis

Three-dimensional GLCM was used to characterize regional spatial intensity patterns, or “texture.” Images were analyzed at 1-mm isotropic resolution. Within each region and sequence, values more than three standard deviations from the regional core mean were excluded. Remaining intensities were divided into 32 gray levels. GLCMs were calculated across 13 spatial directions and averaged before extracting texture features that included entropy, contrast, autocorrelation, and cluster shade. Texture measurements were calculated separately for T1 and FLAIR images and did not require CN reference data. Measurements with insufficient tissue voxels or neighboring voxel pairs were recorded as missing.

### 2.6. Clinical and PET outcomes

Clinical outcomes included the Montreal Cognitive Assessment (MoCA), Mini-Mental State Examination (MMSE), CDR® Dementia Staging Instrument sum-of-boxes score (CDR-SB), and other cohort-available cognitive measures, including the 13-item Alzheimer’s Disease Assessment Scale cognitive subscale and MCSA global cognition. The contemporaneous prediction datasets were restricted to cognitive assessments within 180 days of MRI.

PET comparisons also included tau and amyloid PET results within 180 days of MRI. ¹⁸F-flortaucipir (FTP) PET was used to estimate tau burden. Amyloid PET studies included ¹⁸F-florbetapir (FBP), ¹⁸F-florbetaben (FBB), ¹¹C-Pittsburgh compound B (PiB), and ¹⁸F-NAV4694 (NAV). Temporal meta-region tau standardized uptake value ratios (SUVRs) were used for prediction modeling, with ADNI3 as the training cohort and ADNI4 and SCAN/CLARiTI as the test cohorts. In MCSA, available MMSEcalc scores were MMSE-equivalent values derived from the Short Test of Mental Status.[23] These values were included in association analyses but excluded from primary MMSE prediction testing, which required directly administered MMSE scores.

### 2.7. Association analyses

Associations were examined using multivariable linear regression of rank-transformed measurements, separately within each cohort and in pooled analyses. Pooled analyses used within-cohort ranking and cohort-specific covariate adjustment. These analyses were adjusted for age, sex, education, ICV, and scanner characteristics as covariates. When applicable, additional covariates including regional volume, clinical diagnosis, MRI-PET interval, and PET processing differences were used. Benjamini-Hochberg false discovery rate correction was used when multiple comparisons were made. Adjusted p-values less than 0.05 were considered to be statistically significant. Regional mask processing, calculations of WSRLR and CN-reference, 3D GLCM feature extraction, and association analyses were performed using custom Python scripts with NumPy, SciPy, pandas, and NiBabel. Association samples were determined separately for each regional feature and outcome and included eligible participants from all contributing cohorts. These samples overlapped with the prediction datasets, whereas formal region selection and model development were restricted to ADNI3.

### 2.8. Contemporaneous association prediction model development and testing

Outcome-specific ridge,[24] elastic-net,[25] and histogram gradient-boosting[26] models were implemented using scikit-learn (version 1.9.0).[27] These models and TabPFN-v2[28] were trained using ADNI3 and externally tested without refitting or recalibration in ADNI4, SCAN/CLARiTI, and/or OASIS-3, according to outcome availability. Outcomes included contemporaneous MoCA, MMSE, temporal tau PET burden, and global amyloid Centiloids. Baseline inputs included age, sex, education, log ICV, manufacturer indicators, and 36 log-transformed regional volume-to-ICV ratios. Expanded models added four T1/FLAIR WSRLR measurements and 16 GLCM texture features from the entorhinal cortex and hippocampus, selected exclusively through ADNI3 discovery analyses.

Baseline and expanded classical models were tuned separately using identical fivefold cross-validation splits within ADNI3; TabPFN-v2 used a fixed pretrained configuration. Comparisons used identical participants with complete inputs within each outcome. Performance was assessed using R² and mean absolute error for cognition and amyloid, and sample-size-weighted within-cohort Spearman correlations for tau. Cognitive calibration was evaluated separately by cohort and model. Incremental gains were assessed using paired bootstrap confidence intervals and prediction-swap permutation tests with Holm correction. Secondary area under the receiver operating characteristic curve (AUROC) analyses assessed MoCA <26 and MMSE <24. Detailed eligibility, region-selection, model-development, calibration, and statistical procedures are provided in the Supplementary Methods.

## 3. Results

### 3.1. Cohort characteristics

The study included a total of 2,437 baseline datasets from ADNI3 (658), ADNI4 (549), MCSA (747), SCAN/CLARiTI (408), and OASIS-3 (75). The combined SCAN/CLARiTI group consisted of 325 SCAN and 83 CLARiTI datasets. Among 2,387 datasets with available demographic information, the mean age was 71.2 ± 10.2 years (Table 1). There were 1,290 female participants (54%). Clinical classifications included 1,648 cognitively normal participants, 559 with MCI, 175 with dementia, and 14 with other impairments; diagnosis was unavailable for 41 datasets. Among participants with dementia, 141 had clinically attributed AD dementia: 73 in ADNI3, 45 in ADNI4, and 23 in SCAN/CLARiTI.

**Table 1.**
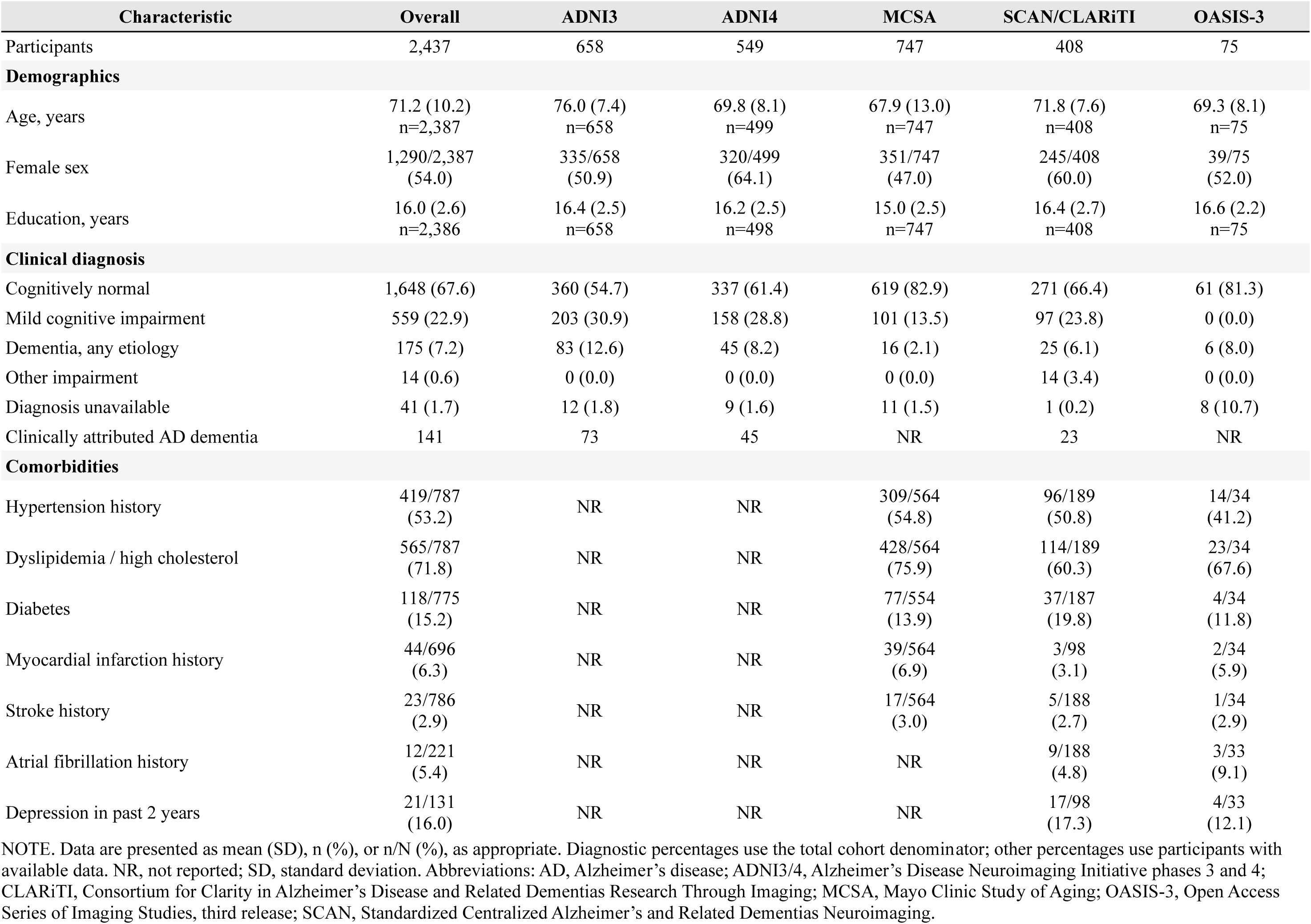
Cohort Characteristics.

### 3.2. Correspondence between WSRLR and CN-referenced signal measurements

For signal measurements, there were moderate to strong correlations between WSRLR and CN-reference normalization methods, with the median regional Spearman correlations ranging from 0.504 to 0.890 (n = 2314; Figure 2B-C, Supplementary Figure 1). For FLAIR, median correlations were 0.504 in ADNI3, 0.718 in ADNI4, 0.856 in MCSA, 0.552 in SCAN, 0.684 in CLARiTI, and 0.817 in OASIS-3. Corresponding correlations for T1 ranged from 0.683 to 0.890. Concordant directions for several principal associations were observed for both normalization methods across matched-participant analyses. For instance, hippocampal FLAIR was negatively associated with local volume using WSRLR and CN referencing (partial r = −0.221 and −0.223, respectively; n = 2,305). Entorhinal FLAIR was negatively associated with regional tau using both methods (partial r = −0.148 and −0.163; n = 587; both q < 0.01).

### 3.3. Regional signal and texture associations with volume and AD dementia

In pooled, covariate-adjusted analyses, WSRLR signal measurements demonstrated statistically significant associations with corresponding regional volume in 15 FLAIR and 20 T1 regions (q<0.05; n=2,376; Figure 3A–B). Higher hippocampal and amygdala FLAIR signals were associated with smaller volumes (r=−0.223 and −0.220), whereas lower entorhinal FLAIR and T1 signals were associated with smaller entorhinal volumes (r=0.092 and 0.144; all q<0.001). GLCM entropy was associated with volume in 18 FLAIR and 24 T1 regions, with predominantly negative associations but with positive hippocampal associations (Figure 3C-D). Other texture features also showed widespread volume associations (Supplementary Figure 2). Notably, contrast and autocorrelation showed generally negative correlations with regional volume, while cluster shade showed positive correlations. Sensitivity analyses with additional adjustment for clinical diagnosis retained most signal-volume and entropy-volume associations, but hippocampal FLAIR entropy was no longer significant.

**Figure 3.**
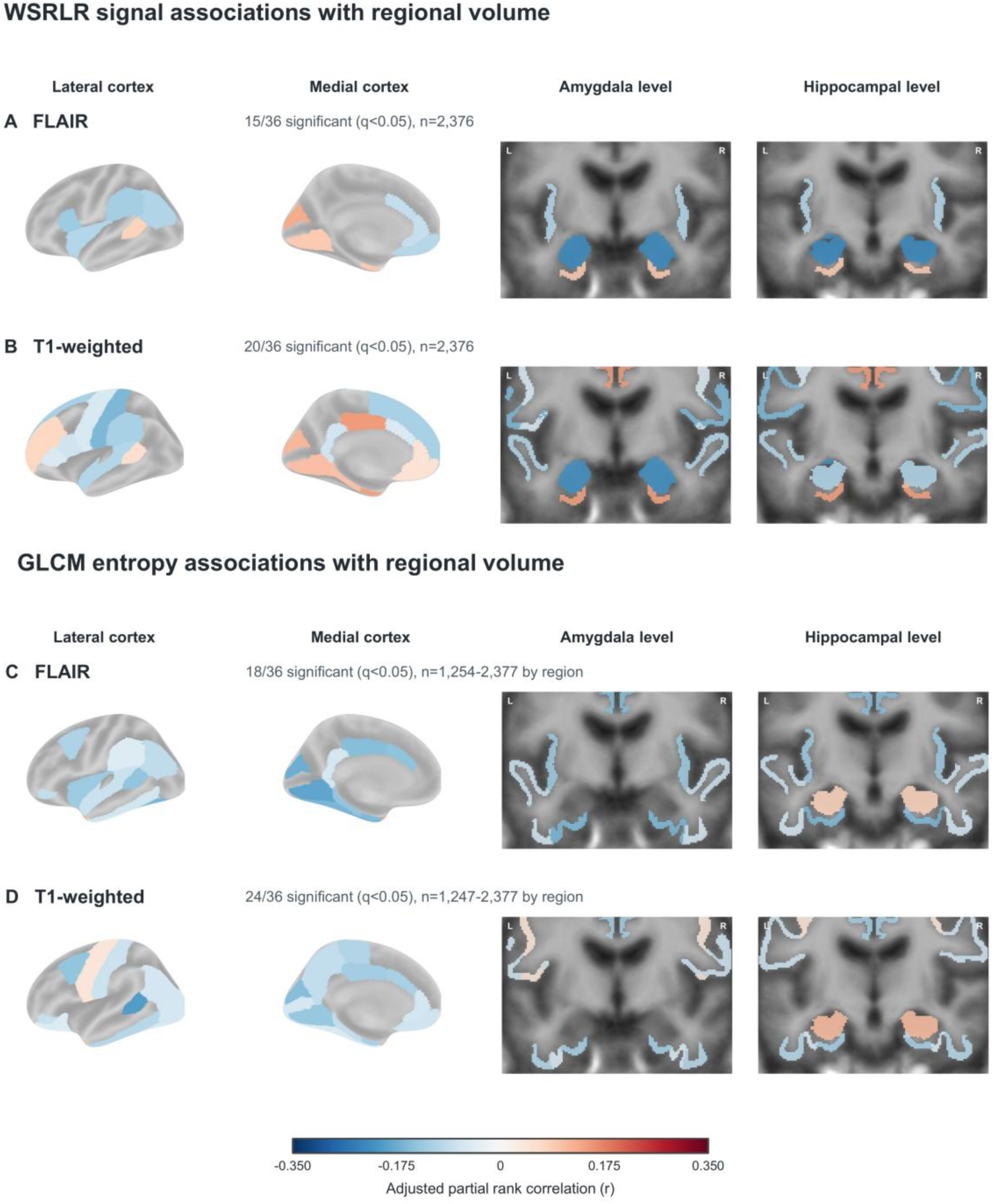
Regional signal and texture (entropy) associations with corresponding brain volume. FLAIR/T1 maps show pooled, cohort-stratified partial rank correlations of WSRLR (A/B) and GLCM entropy (C/D) with regional volume/ICV. Covariates were age, sex, education, ICV, and scanner. Colored regions survived Benjamini-Hochberg FDR correction (q<0.05): 15/20 for WSRLR and 18/24 for entropy (FLAIR/T1). Red indicates positive and blue negative associations. Sample sizes represent unique participants per regional analysis: WSRLR, n=2,376 for both sequences; entropy, n=1,254–2,377 for FLAIR and n=1,247–2,377 for T1. Abbreviations: FDR, false discovery rate; FLAIR, fluid-attenuated inversion recovery; GLCM, gray-level co-occurrence matrix; ICV, intracranial volume; WSRLR, within-scan regional-to-rest log ratio.

After adjusting for regional volume and other covariates, clinically attributed AD dementia was associated with lower entorhinal FLAIR WSRLR (r=−0.192; n=1,085, Supplementary Figure 3A) and greater entorhinal FLAIR entropy (r=0.224; n=1,086; both q<0.001, Supplementary Figure 3B), compared to CN. Conversely, AD dementia was associated with higher hippocampal and amygdala FLAIR WSRLR (r=0.237 and 0.260, respectively) and lower hippocampal FLAIR entropy (r=−0.152; all q<0.001). Additional brain regions that demonstrated significantly different correlations between AD dementia and CN participants are described in Supplementary Figure 4.

### 3.4. Cognitive and tau PET associations beyond regional volume

Higher hippocampal FLAIR WSRLR was associated with lower MoCA scores when adjusted for corresponding regional volume and other covariates (r=−0.162; q<0.001; n=1,506, Figure 4A). Greater entorhinal FLAIR entropy was associated with lower MoCA and MMSE scores (r=−0.198 and −0.098, respectively; both q<0.001; Supplementary Figure 5). In general, T1 texture showed broader associations with cognitive performance (Figure 4C-F). Notably, greater isthmus-cingulate entropy was correlated with lower MoCA and MMSE scores (r=−0.233 and −0.206), while hippocampal cluster shade correlated with higher scores (r=0.224 and 0.191; all q<0.001; MoCA n=1,507; MMSE n=1,944; Figure 4). Additional adjustment for clinical diagnosis attenuated cognitive associations, although entorhinal FLAIR entropy remained associated with lower MoCA scores (r=−0.132; q<0.001; n=1,485). Selected T1 texture associations also persisted, including isthmus-cingulate entropy and hippocampal cluster shade; the latter was associated with both MoCA and MMSE scores (r=0.103 and 0.090, respectively; both q=0.008).

**Figure 4.**
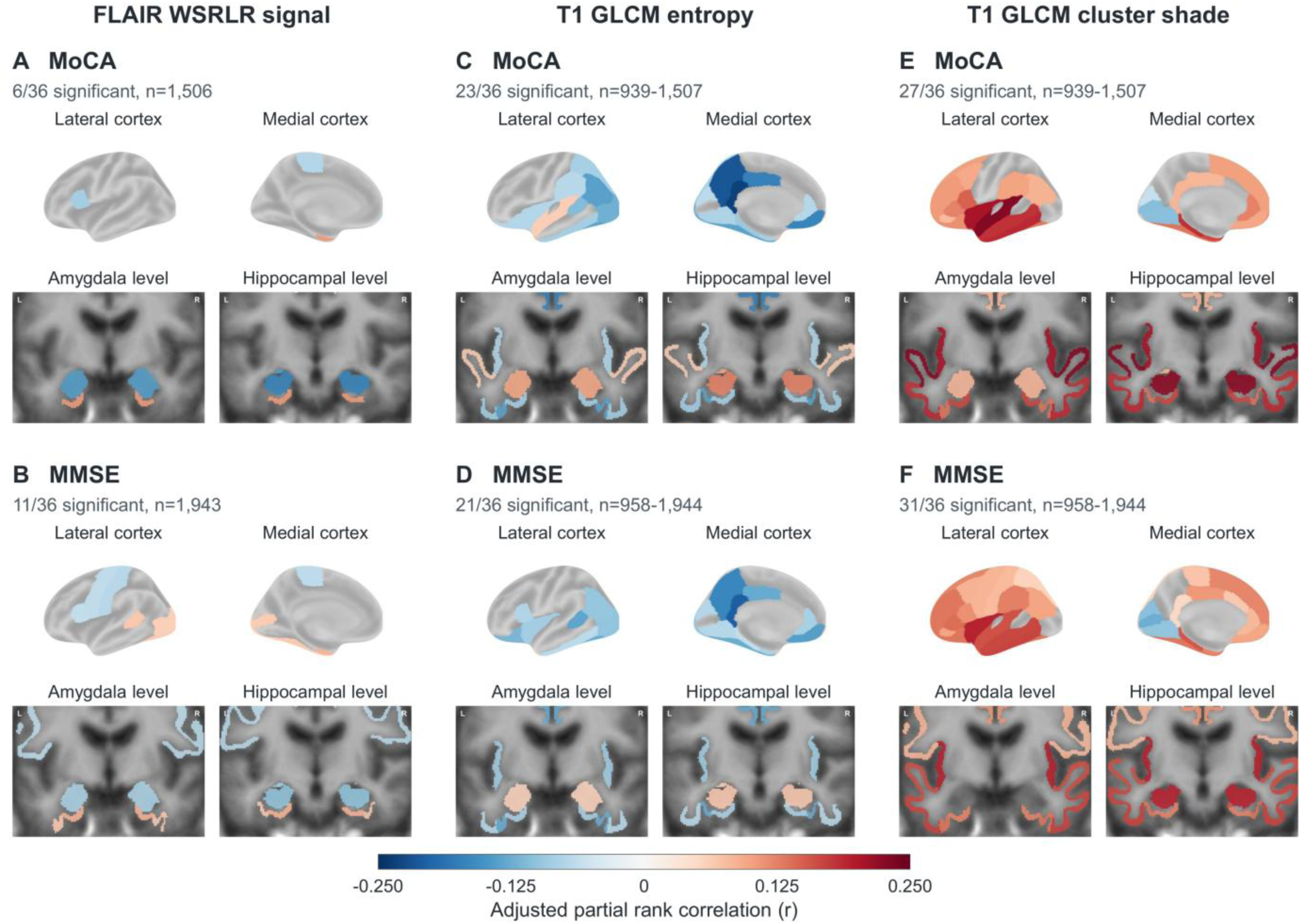
Regional signal and texture associations with cognitive performance. Pooled partial rank correlations relate MoCA (A, C, E) and MMSE (B, D, F) to FLAIR WSRLR (A, B), T1 GLCM entropy (C, D), and T1 cluster shade (E, F). Models adjusted for regional volume and other covariates, without clinical diagnosis. Significant region counts for MoCA/MMSE were 6/11, 23/21, and 27/31, respectively. Only Benjamini-Hochberg FDR-adjusted q<0.05 associations are colored; red indicates higher and blue lower cognitive scores with increasing feature values. Sample sizes represent unique participants per regional analysis: FLAIR WSRLR, n=1,506 for MoCA and n=1,943 for MMSE; T1 texture, n=939–1,507 for MoCA and n=958–1,944 for MMSE. Abbreviations: FDR, false discovery rate; FLAIR, fluid-attenuated inversion recovery; GLCM, gray-level co-occurrence matrix; MMSE, Mini-Mental State Examination; MoCA, Montreal Cognitive Assessment; WSRLR, within-scan regional-to-rest log ratio.

Correlational analyses between PET and signal/texture measurements revealed directionally concordant associations in both tau and amyloid uptake in a subset of brain regions. Higher T1 WSRLR in the postcentral and superior-parietal cortices, as well as greater paracentral T1 entropy and contrast, were associated with higher corresponding regional PET uptake (tau: r=0.124–0.175, n=607; amyloid: r=0.102–0.159, n=924; all q<0.05; Figure 5A). Entorhinal associations were more evident for tau; lower FLAIR signal and lower T1 cluster shade were associated with higher regional tau uptake (r=−0.146, q=0.010 and r=−0.205, q<0.001, respectively), but the corresponding amyloid associations were not statistically significant after FDR correction (Figure 5B).

**Figure 5.**
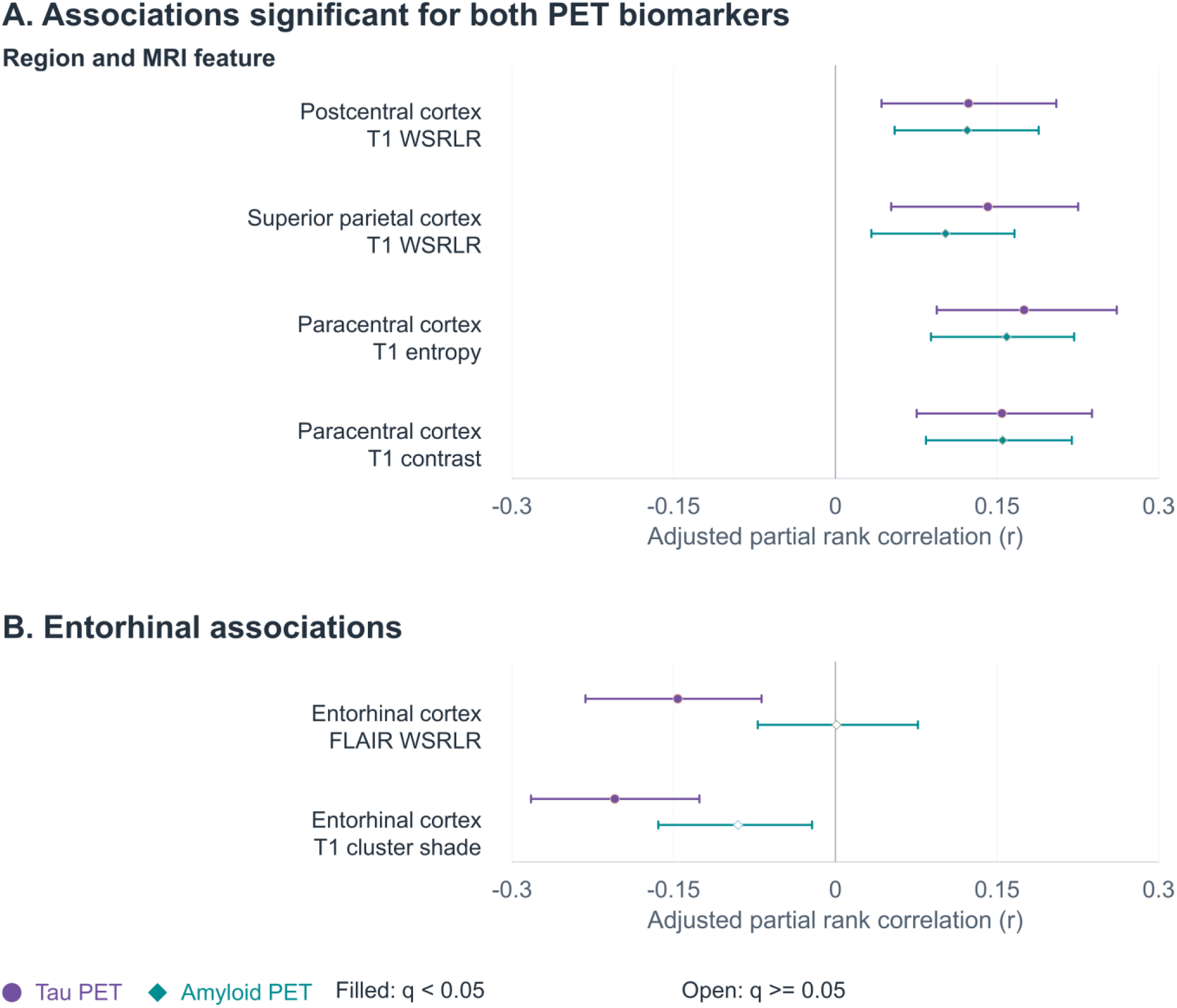
Regional signal and texture associations with tau and amyloid PET beyond volume. (A) Selected T1 WSRLR, entropy, and contrast associations significant for both biomarkers. (B) Entorhinal FLAIR WSRLR and T1 cluster shade associations. Points are partial rank correlations adjusted for regional volume, diagnosis, MRI-PET interval, and other covariates. Filled markers indicate Benjamini-Hochberg FDR-adjusted q<0.05; open markers indicate nonsignificance. Bars show nominal 95% confidence intervals from 2,000 cohort-stratified participant bootstraps, with re-ranking and nuisance-model refitting. Intervals are not multiplicity-adjusted and may exclude zero despite a nonsignificant q-value. Sample sizes represent unique participants per association: tau, n=607, and amyloid, n=924, except entorhinal T1 cluster shade (n=608 and n=925, respectively). Abbreviations: FDR, false discovery rate; FLAIR, fluid-attenuated inversion recovery; MRI, magnetic resonance imaging; PET, positron emission tomography; WSRLR, within-scan regional-to-rest log ratio.

### 3.5. Incremental gains for predicting contemporaneous cognition and amyloid/tau burden

To evaluate incremental gains of signal and texture for the prediction of contemporaneous cognitive performance and amyloid/tau burden, ADNI3-trained ridge, elastic-net, histogram gradient-boosting, and TabPFN-v2 models were externally tested in ADNI4, SCAN/CLARiTI, and/or OASIS-3, comparing demographic and volumetric inputs with models additionally incorporating entorhinal and hippocampal signal and texture. Incomplete inputs or processing ineligibility excluded 36 participants from MoCA, 52 from MMSE, and one from each PET analysis, all from ADNI4. Combined signal and texture improved pooled MoCA prediction with ridge, elastic-net, and gradient-boosting models (n=868; ΔR²=0.076–0.104; all Holm-adjusted P<0.002), and MMSE prediction with elastic-net, gradient-boosting, and TabPFN-v2 models (n=551; ΔR²=0.080–0.157; all adjusted P<0.05; Table 2). All four models showed higher MoCA R² and lower MAE in each testing cohort, although confidence intervals included zero improvement in OASIS-3. MMSE gains were consistent across models in ADNI4 but varied in OASIS-3 (Supplementary Table 3). Calibration slopes approached the ideal value across all cohort–model comparisons, although systematic overestimation or underestimation did not consistently decrease (Supplementary Table 4). Adding signal and texture also improved discrimination of MoCA <26 in three models (ΔAUROC=0.026–0.029) and MMSE <24 in two models (ΔAUROC=0.036–0.093; all Holm-adjusted P≤0.005; Supplementary Table 5).

**Table 2.**
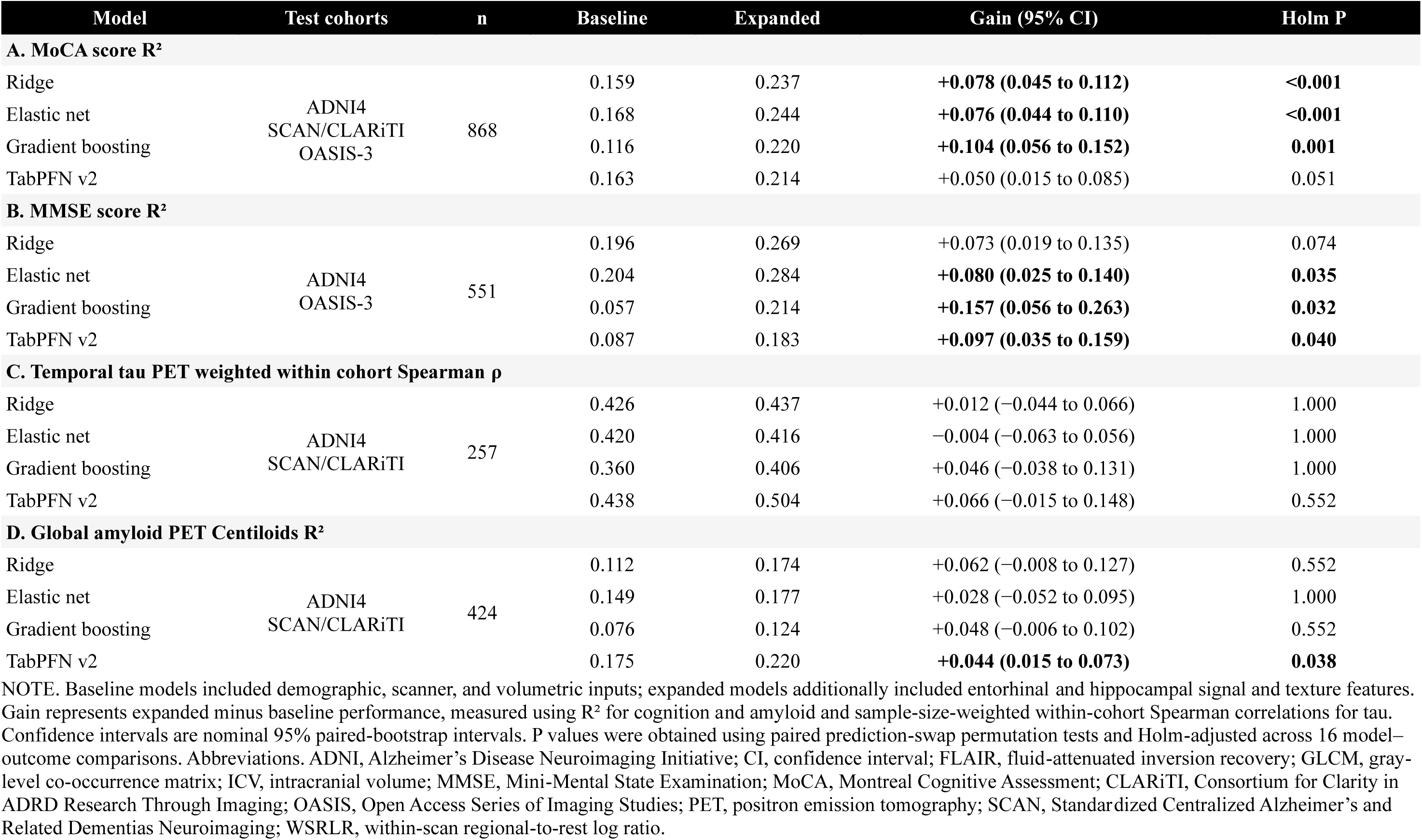
Incremental prediction gains of hippocampal and entorhinal signal and texture models for prediction of contemporaneous cognitive performance and PET.

WSRLR alone improved ridge and elastic-net predictions of MoCA (ΔR²=0.024–0.028) and MMSE (ΔR²=0.033–0.035; all adjusted P≤0.005). Adding WSRLR to texture-containing models further improved MoCA prediction (ΔR²=0.013–0.017; both adjusted P<0.02), but not MMSE prediction (Supplementary Table 2). To assess whether predictive gains were specific to the selected medial temporal regions, we evaluated pericalcarine and pericalcarine–cuneus panels as anatomical negative-control comparisons. Neither pericalcarine nor pericalcarine–cuneus comparison panels significantly improved cognitive prediction; the entorhinal–hippocampal panel outperformed the feature-count-matched pericalcarine–cuneus panel for MoCA (ΔR²=0.071; 95% CI, 0.038–0.109; adjusted P<0.001) and MMSE (ΔR²=0.090; 95% CI, 0.033–0.154; adjusted P=0.014). PET prediction gains were less consistent: no model significantly improved temporal tau estimation (n=257; all adjusted P≥0.552), while only TabPFN-v2 improved amyloid Centiloid estimation (n=424; R²=0.175 to 0.220; ΔR²=0.044; 95% CI, 0.015–0.073; adjusted P=0.038; Table 2).

## 4. Discussion

### 4.1. Within-scan characterization

Our findings support WSRLR as a practical approach for characterizing relative regional MRI signal without scanner-specific CN reference data. Previous studies have demonstrated the biological relevance of normalized signal intensity measurements from conventional MRI. Existing normalization strategies have utilized comparisons against normative references derived from an external cohort[6] or cross-sequence intensity ratios such as T1-/T2-weighted imaging.[29] Previous within-scan normalization efforts have used pre-defined anatomic reference regions.[11,12] WSRLR instead characterizes the regional intensity profile within each image contrast using a distributed regional reference, which permits regional contrast assessments without a separate normative population, cross-sequence references, or specific, designated anatomical structures. Moderate-to-strong correlations observed with CN-referenced measurements, associations with AD-related pathologies, and contributions to contemporaneous predictions support their value in within-scan characterization of regional signal changes. However, these measurements remain relative and are subject to within-scan variations in imaging due to artifacts and magnetic inhomogeneity. Future studies comparing WSRLR measurements with quantitative T1 and T2 relaxation times obtained using magnetic resonance fingerprinting[30] may help validate WSRLR and clarify AD-related associations.

### 4.2. Complementary regional signal and texture findings

Regional signal measurements demonstrated differing associations with AD-related abnormalities. Notably, lower entorhinal FLAIR WSRLR was associated with smaller corresponding regional volume, clinically attributed AD dementia, and greater regional tau uptake. Conversely, higher hippocampal and amygdala FLAIR WSRLR measurements were associated with smaller volumes and AD dementia. The entorhinal findings are broadly in line with observations from the DIAN study that demonstrated lower cortical FLAIR signal to greater AD pathology.[6] In comparison, a separate study showed higher left-to-right piriform-amygdala FLAIR intensity ratios associated with greater medial-temporal and cerebral atrophy.[31] Regional FLAIR differences were also observed in neurologically normal individuals, including relatively high hippocampal and amygdala signal.[32] These findings suggest that relative FLAIR intensity should not be interpreted as a uniform measure of tissue injury. Rather, these may reflect differences in regional tissue composition, disease-related alterations, and possibly underlying compensatory mechanisms rather than a single underlying pathophysiological process.

Similarly, texture-measurements demonstrated region-and feature-dependent associations. Greater entorhinal FLAIR entropy was associated with smaller regional volume, AD dementia, and poorer cognition, while hippocampal entropy was lower in AD dementia. These observations complement previous T1-based findings linking hippocampal texture to cognition and glucose metabolism independently of volume, and identifying entorhinal texture differences across cognitively normal, MCI, and AD groups.[33,34] Our PET analyses further demonstrated that lower entorhinal T1 cluster shade was associated with greater regional tau uptake, while greater paracentral T1 entropy and contrast were associated with both tau and amyloid uptake after adjusting for volume and diagnosis. These feature-dependent patterns are broadly in alignment with prior findings by Lee and Kim linking higher cortical T1 entropy and contrast, but lower hippocampal and entorhinal cluster shade, to greater tau burden.[8] Unlike the previous study, which found no significant amyloid associations,[8] our results suggest regional texture-amyloid relationships. This may be attributed to the differences in cohort (ADNI3 in the previous study vs. multi-cohort design in our study) and differences in MRI/PET acquisitions. Overall, our findings extend this predominantly T1-based literature through combined T1 and FLAIR assessment and volume-adjusted associations with clinical and PET outcomes.

### 4.3. Incremental predictive value and clinical implications

Adding entorhinal and hippocampal signal and texture measurements to demographic and whole-brain regional volumetric inputs improved estimation of contemporaneous cognitive performance. In pooled external testing, MoCA gains were positive across all four modeling strategies and statistically significant in three. MMSE estimation also improved significantly in three out of four models, suggesting that the improved prediction was not restricted to a specific algorithm. These findings complement previous findings that hippocampal texture adds information beyond volume for predicting cognitive changes,[9] although our models examined contemporaneous rather than future outcomes. Improvements were less evident for the prediction of tau or amyloid burden, however, which suggests that associations with regional PET uptake do not necessarily translate into proven prediction of an individual’s overall PET burden.

Clinically, our results suggest that these measurements may potentially support opportunistic identification of patients who warrant formal cognitive assessment when conventional MRI is already available. WSRLR allows relative regional signal assessment within a particular sequence without the need for scanner-specific CN references, cross-sequence calculations, or a reliance on a single anatomical reference region. GLCM texture features also allow characterization of within-sequence spatial intensity patterns without requiring normative reference data. However, absolute predictive performance remained moderate overall, indicating that these measurements would require further validation, and that their potential role may be to prompt further cognitive evaluation and, when appropriate, AD biomarker testing. Early recognition of early AD is increasingly relevant as anti-amyloid therapies become available for appropriately selected patients with confirmed amyloid pathology.[35,36] While the present findings do not establish screening accuracy or treatment eligibility, future prospective studies may help determine appropriate imaging thresholds that warrant further cognitive/AD evaluation and assess clinical benefits.

### 4.4. Limitations and future directions

There are several limitations to our study. This is a retrospective, predominantly cross-sectional study that cannot establish causality. It is thus not feasible to determine whether signal and texture abnormalities precede atrophy or cognitive decline. The cohorts differed in terms of participant characteristics, diagnostic evaluations, and imaging protocols. While covariates were adjusted as applicable for different hypotheses, there may be residual confounding that is not eliminated. Clinical AD diagnoses were also not uniformly biomarker-confirmed, and differences in PET availability, processing methods, and MRI-PET intervals limit cross-cohort comparability. Furthermore, the research cohorts may not reflect individuals undergoing routine clinical MRI, limiting our perspectives about opportunistic screening. Prediction analyses required complete demographic and imaging inputs. This may introduce selection bias and limit generalizability, as excluded ADNI4 participants had lower median MoCA (22 versus 25) and MMSE (28 versus 29) scores than included participants. Residual partial-volume effects and segmentation errors may influence regional measurements despite tissue-core masking and adjustment for regional volume, particularly in small or atrophic structures such as the entorhinal cortex. Lastly, WSRLR characterizes relative rather than absolute signal, and widespread changes across the entire brain can attenuate its sensitivity for relative regional signal changes. Future studies comparing WSRLR against quantitative MRI such as MR fingerprinting or histopathology may help further validate this approach. Longitudinal studies are also needed to determine the value of within-scan analyses for predicting future regional atrophy, amyloid/tau burden, cognitive decline and/or AD diagnosis.

## 5. Conclusions

WSRLR allowed within-scan characterization of relative T1 and FLAIR signal without scanner-specific CN reference populations. Regional signal and texture measurements exhibited distinct associations with volume, AD dementia, cognitive performance, and PET biomarkers, including selected associations beyond regional volume. Adding signal and texture to demographic and volumetric inputs improved prediction of contemporaneous cognitive performance; incremental prediction gain for contemporaneous amyloid and tau PET was less consistent. These findings suggest that our within-scan approach may allow for opportunistic analyses of additional biomarkers that complement volumetry from conventional MRI and support prospective validation to examine its clinical screening utility and prediction of future decline.

## Acknowledgments

We thank the participants, their families and study partners, investigators, and staff of ADNI, the Mayo Clinic Study of Aging, NACC and its contributing Alzheimer’s Disease Research Centers, SCAN, CLARiTI, and OASIS-3 for making this research possible.

Support for ADNI data collection and sharing is provided by National Institute on Aging award U19AG024904 to the Northern California Institute for Research and Education. Previous support included the National Institute of Biomedical Imaging and Bioengineering, Canadian Institutes of Health Research, and private contributions facilitated through the Foundation for the National Institutes of Health from AbbVie; Alzheimer’s Association; Alzheimer’s Drug Discovery Foundation; Araclon Biotech; BioClinica, Inc.; Biogen; Bristol-Myers Squibb Company; CereSpir, Inc.; Cogstate; Eisai Inc.; Elan Pharmaceuticals, Inc.; Eli Lilly and Company; EuroImmun; F. Hoffmann-La Roche Ltd and Genentech, Inc.; Fujirebio; GE Healthcare; IXICO Ltd.; Janssen Alzheimer Immunotherapy Research & Development, LLC; Johnson & Johnson Pharmaceutical Research & Development LLC; Lumosity; Lundbeck; Merck & Co., Inc.; Meso Scale Diagnostics, LLC; NeuroRx Research; Neurotrack Technologies; Novartis Pharmaceuticals Corporation; Pfizer Inc.; Piramal Imaging; Servier; Takeda Pharmaceutical Company; and Transition Therapeutics.

Mayo Clinic Study of Aging data were collected with National Institutes of Health support under U01 AG006786, led by Ronald C. Petersen, MD, PhD.

The NACC database is funded by NIA/NIH Grant U24 AG072122.

SCAN is a multi-institutional project that was funded as a U24 grant (AG067418) by the National Institute on Aging in May 2020. Data collected by SCAN and shared by NACC are contributed by the NIA-funded ADRCs as follows:

Arizona Alzheimer’s Center - P30 AG072980 (PI: Eric Reiman, MD); R01 AG069453 (PI: Eric Reiman (contact), MD); P30 AG019610 (PI: Eric Reiman, MD); and the State of Arizona which provided additional funding supporting our center; Boston University - P30 AG013846 (PI Neil Kowall MD); Cleveland ADRC - P30 AG062428 (James Leverenz, MD); Cleveland Clinic, Las Vegas - P20AG068053; Columbia - P50 AG008702 (PI Scott Small MD); Duke/UNC ADRC - P30 AG072958; Emory University - P30AG066511 (PI Levey Allan, MD, PhD); Indiana University - R01 AG19771 (PI Andrew Saykin, PsyD); P30 AG10133 (PI Andrew Saykin, PsyD); P30 AG072976 (PI Andrew Saykin, PsyD); R01 AG061788 (PI Shannon Risacher, PhD); R01 AG053993 (PI Yu-Chien Wu, MD, PhD); U01 AG057195 (PI Liana Apostolova, MD); U19 AG063911 (PI Bradley Boeve, MD); and the Indiana University Department of Radiology and Imaging Sciences; Johns Hopkins - P30 AG066507 (PI Marilyn Albert, Phd.); Mayo Clinic - P50 AG016574 (PI Ronald Petersen MD PhD); Mount Sinai - P30 AG066514 (PI Mary Sano, PhD); R01 AG054110 (PI Trey Hedden, PhD); R01 AG053509 (PI Trey Hedden, PhD); New York University - P30AG066512-01S2 (PI Thomas Wisniewski, MD); R01AG056031 (PI Ricardo Osorio, MD); R01AG056531 (PIs Ricardo Osorio, MD; Girardin Jean-Louis, PhD); Northwestern University - P30 AG013854 (PI Robert Vassar PhD); R01 AG045571 (PI Emily Rogalski, PhD); R56 AG045571, (PI Emily Rogalski, PhD); R01 AG067781, (PI Emily Rogalski, PhD); U19 AG073153, (PI Emily Rogalski, PhD); R01 DC008552, (M.-Marsel Mesulam, MD); R01 AG077444, (PIs M.-Marsel Mesulam, MD, Emily Rogalski, PhD); R01 NS075075 (PI Emily Rogalski, PhD); R01 AG056258 (PI Emily Rogalski, PhD); Oregon Health and Science University - P30 AG008017 (PI Jeffrey Kaye MD); R56 AG074321 (PI Jeffrey Kaye, MD); Rush University - P30 AG010161 (PI David Bennett MD); Stanford - P30AG066515; P50 AG047366 (PI Victor Henderson MD MS); University of Alabama, Birmingham - P20; University of California, Davis - P30 AG10129 (PI Charles DeCarli, MD); P30 AG072972 (PI Charles DeCarli, MD); University of California, Irvine - P50 AG016573 (PI Frank LaFerla PhD); University of California, San Diego - P30AG062429 (PI James Brewer, MD, PhD); University of California, San Francisco - P30 AG062422 (Rabinovici, Gil D., MD); University of Kansas - P30 AG035982 (Russell Swerdlow, MD); University of Kentucky - P30 AG028283-15S1 (PIs Linda Van Eldik, PhD and Brian Gold, PhD); University of Michigan ADRC - P30AG053760 (PI Henry Paulson, MD, PhD) P30AG072931 (PI Henry Paulson, MD, PhD) Cure Alzheimer’s Fund 200775 - (PI Henry Paulson, MD, PhD) U19 NS120384 (PI Charles DeCarli, MD, University of Michigan Site PI Henry Paulson, MD, PhD) R01 AG068338 (MPI Bruno Giordani, PhD, Carol Persad, PhD, Yi Murphey, PhD) S10OD026738-01 (PI Douglas Noll, PhD) R01 AG058724 (PI Benjamin Hampstead, PhD) R35 AG072262 (PI Benjamin Hampstead, PhD) W81XWH2110743 (PI Benjamin Hampstead, PhD) R01 AG073235 (PI Nancy Chiaravalloti, University of Michigan Site PI Benjamin Hampstead, PhD) 1I01RX001534 (PI Benjamin Hampstead, PhD) IRX001381 (PI Benjamin Hampstead, PhD); University of New Mexico - P20 AG068077 (Gary Rosenberg, MD); University of Pennsylvania - State of PA project 2019NF4100087335 (PI David Wolk, MD); Rooney Family Research Fund (PI David Wolk, MD); R01 AG055005 (PI David Wolk, MD); University of Pittsburgh - P50 AG005133 (PI Oscar Lopez MD); University of Southern California - P50 AG005142 (PI Helena Chui MD); University of Washington - P50 AG005136 (PI Thomas Grabowski MD); University of Wisconsin - P50 AG033514 (PI Sanjay Asthana MD FRCP); Vanderbilt University - P20 AG068082; Wake Forest - P30AG072947 (PI Suzanne Craft, PhD); Washington University, St. Louis - P01 AG03991 (PI John Morris MD); P01 AG026276 (PI John Morris MD); P20 MH071616 (PI Dan Marcus); P30 AG066444 (PI John Morris MD); P30 NS098577 (PI Dan Marcus); R01 AG021910 (PI Randy Buckner); R01 AG043434 (PI Catherine Roe); R01 EB009352 (PI Dan Marcus); UL1 TR000448 (PI Brad Evanoff); U24 RR021382 (PI Bruce Rosen); Avid Radiopharmaceuticals / Eli Lilly; Yale - P50 AG047270 (PI Stephen Strittmatter MD PhD); R01AG052560 (MPI: Christopher van Dyck, MD; Richard Carson, PhD); R01AG062276 (PI: Christopher van Dyck, MD); 1Florida - P30AG066506-03 (PI Glenn Smith, PhD); P50 AG047266 (PI Todd Golde MD PhD)

This research was supported by the ADRC Consortium for Clarity in ADRD Research Through Imaging (CLARiTI) with funding from the National Institute on Aging (NIA) U01AG082350.

The NACC database is funded by NIA/NIH Grant U24 AG072122. SCAN is a multi-institutional project that was funded as a U24 grant (AG067418) by the National Institute on Aging in May 2020. CLARiTI data are contributed by the NIA-funded ADRCs at the following link: https://www.naccdata.org/publish-with-nacc-data/.

Data were provided in part by the Open Access Series of Imaging Studies (OASIS). The OASIS-3 Longitudinal Multimodal Neuroimaging resource was led by T. Benzinger, D. Marcus, and J. Morris, with support from NIH awards P30 AG066444, P50 AG00561, P30 NS09857781, P01 AG026276, P01 AG003991, R01 AG043434, UL1 TR000448, and R01 EB009352. OASIS-3_AV1451 was led by T. Benzinger and J. Morris, with support from P30 AG066444 and AW00006993. Avid Radiopharmaceuticals, a wholly owned subsidiary of Eli Lilly, provided AV-45 and AV-1451 tracer doses for the contributing datasets.

## Use of generative AI

ChatGPT and Codex (OpenAI) assisted with literature searches, analysis-code development and debugging, figure preparation, and manuscript drafting and editing. The authors reviewed and edited the AI-assisted material and take responsibility for the analyses, interpretation, and final content.

## Consent Statement

This study used existing de-identified data from ADNI, MCSA, SCAN/CLARiTI, and OASIS-3 under their respective data-use agreements. Ethics approval and informed consent for the original data collection were managed by the contributing studies according to their respective protocols. No participants were recruited or contacted for the present secondary analysis.

## Data availability

Data used in this study were obtained from the Alzheimer’s Disease Neuroimaging Initiative, Mayo Clinic Study of Aging, National Alzheimer’s Coordinating Center, including SCAN and CLARiTI, and OASIS-3. Access is available through the respective data providers, subject to approval and applicable data-use agreements. Participant-level source and derived data are not publicly redistributed by the authors because of these agreements. Aggregate results are reported in the manuscript and supplementary material.

## Funding

This work was funded by the Veterans Affairs Early Career Development program (b.c.y). The funder had no role in the study design, data analysis or interpretation, manuscript preparation, or the decision to submit the manuscript for publication.

## Declaration of interest

The authors declare no conflicts of interest.

## Author contributions

Wontack Han: Methodology, formal analysis, and writing—review and editing. Hyun-Jee Sarah Han: Methodology, formal analysis, and writing—review and editing. Dong-Joo Kim: Resources and writing—review and editing. Byung Chul Yoon: Conceptualization, data curation, formal analysis, and writing—original draft.

## Supplementary Materials

Supplementary Methods. Contemporaneous prediction analyses

Supplementary Table 1. Brain regions included in the MRI analyses

Supplementary Table 2. Contributions of WSRLR and texture to external cognitive prediction

Supplementary Table 3. Cohort-specific cognitive prediction performance

Supplementary Table 4. Calibration of external cognitive predictions

Supplementary Table 5. Discrimination of low cognitive scores after adding MRI signal and texture

Supplementary Figure 1. Correlations between WSRLR and CN-referenced T1-weighted and FLAIR signal measurements across cohorts

Supplementary Figure 2. Regional GLCM contrast, autocorrelation, and cluster-shade associations with corresponding brain volume

Supplementary Figure 3. Representative entorhinal FLAIR signal and texture differences between cognitively normal participants and participants with clinically attributed AD dementia

Supplementary Figure 4. Regional WSRLR signal and GLCM texture differences between clinically attributed AD dementia and cognitively normal participants

Supplementary Figure 5. Selected regional signal and texture associations with cognitive performance beyond regional volume

## Notes

### Competing Interest Statement

The authors have declared no competing interest.

