## SupplementaryMaterial for "Within-scan MRI signal and texture in Alzheimer’s disease: a multicohort study"

**Supplementary Methods: Contemporaneous prediction analyses**

### S1. Prediction samples and eligibility

Outcome-specific models were developed using ADNI3 and evaluated in the external cohorts listed below. Cognitive and PET outcomes were matched to MRI within 180 days. Participants required complete demographic and volumetric predictors, valid T1 and FLAIR WSRLR profiles across the 36-region panel, and complete entorhinal and hippocampal texture measurements. Outcomes were not imputed. Baseline and expanded models used identical participants within each outcome, and these samples were shared across algorithms. MCSA was excluded from direct MMSE prediction because its available MMSE-equivalent values were derived from the Short Test of Mental Status rather than direct MMSE administration.

| Outcome | ADNI3 training | ADNI4 testing | NACC testing | OASIS-3 testing |
|---|---|---|---|---|
| MoCA | 639 | 422 | 380 | 66 |
| MMSE | 650 | 485 | — | 66 |
| Temporal tau PET | 351 | 198 | 59 | — |
| Global amyloid Centiloids | 469 | 319 | 105 | — |

### S2. Candidate-region selection and predictor panels

Candidate regions were selected exclusively from ADNI3 association analyses. Each region required at least one T1/FLAIR signal or texture association with FDR-adjusted $q<0.05$ in each of four domains: clinically attributed AD dementia versus CN, corresponding regional volume, cognitive performance, and corresponding regional tau or amyloid uptake. Different features could qualify different domains. PET qualification used diagnosis-adjusted associations, whereas the other domains used the primary association models.

This procedure retained the entorhinal cortex and hippocampus. Selection used the earlier cortical-reference WSRLR implementation; the selected panel remained unchanged after adopting the 36-region reference. All four WSRLR measurements and 16 texture features from these regions were retained across algorithms and outcomes. External cohorts were not used in the formal region-selection procedure.

The baseline panel contained 43 predictors: age, sex, education, log ICV, three manufacturer indicators, and 36 log-transformed regional volume-to-ICV ratios. The combined expanded panel contained 63 predictors. Additional cognitive analyses evaluated signal alone, texture alone, and the contribution of signal beyond texture.

### S3. Model implementation and tuning

Ridge, elastic-net, and histogram gradient-boosting regression models were implemented using scikit-learn version 1.9.0. Hyperparameters were selected separately for each outcome, algorithm, and predictor panel using identical shuffled fivefold splits within ADNI3, minimizing cross-validated mean squared error. Predictor scaling used RobustScaler and was fitted within each training fold. Elastic-net outcomes were additionally standardized within the training pipeline. The tuning grids were as follows:

| Model | Hyperparameters examined |
|---|---|
| Ridge | Regularization strength α: 1, 10, 100, 1000 |

| Model | Hyperparameters examined |
|---|---|
| Elastic net | α: 0.001, 0.01, 0.1, 1; L1 mixing proportion: 0.1, 0.5, 0.9 |
| Histogram gradient boosting | Maximum leaf nodes: 7 or 15; L2 regularization: 10 or 30 |

Elastic-net fitting allowed 20,000 iterations with a convergence tolerance of $10^{-6}$. Gradient boosting used 200 iterations, a learning rate of 0.05, a minimum leaf size of 20, and no early stopping. Cross-validation used a fixed random state of 20261114.

TabPFN-v2 was implemented using package version 2.2.1 and pretrained v2 regression weights, with four estimators and a random state of 20261015. Its configuration was fixed without hyperparameter tuning.

Following development, models were fitted using all eligible ADNI3 participants and applied externally without refitting or recalibration. Cognitive predictions were bounded to 0–30 and tau predictions to nonnegative values; negative amyloid Centiloids were retained.

**S4. Performance, calibration, and statistical inference**

Cognitive and amyloid prediction performance was evaluated using $R^2$ and mean absolute error, both within individual cohorts and in pooled external samples. Because tau processing and measurement scales differed across cohorts, pooled tau performance used the sample-size-weighted mean of within-cohort Spearman correlations.

Cognitive calibration was assessed separately for each cohort, algorithm, and predictor panel by regressing observed scores on predicted scores:

$$\text{Observed score} = \alpha + \beta \times \text{Predicted score}.$$

Ideal calibration corresponds to an intercept of 0 and slope of 1. Mean prediction error was calculated as predicted minus observed score; positive values indicate overestimation. Calibration regressions were diagnostic and were not used to modify predictions.

Confidence intervals used 5,000 paired participant bootstrap resamples. Pooled resampling was stratified by cohort, preserving cohort sample sizes. Baseline and expanded predictions remained paired within participants. Calibration intervals were calculated within each cohort. Reported 95% intervals were percentile intervals and were not multiplicity-adjusted.

Two-sided P values for incremental performance were obtained from 20,000 permutations that randomly exchanged baseline and expanded predictions within participants. Holm correction was applied separately across 16 primary model–outcome comparisons and 32 cognitive signal-versus-texture comparisons. These uncertainty estimates were conditional on the fitted models and did not incorporate variability from repeating model development. Exploratory discrimination analyses evaluated MoCA $<26$ and MMSE $<24$ using negative continuous predictions from the frozen models, without refitting or recalibration. AUROC gains were assessed using the paired bootstrap and prediction-swap procedures described above, with separate Holm correction across four models for each outcome (Supplementary Table 5).

**S5. Anatomical-comparison analyses**

Exploratory ridge analyses compared the entorhinal–hippocampal panel with pericalcarine features alone and a pericalcarine–cuneus panel. Pericalcarine features added two WSRLR and eight texture measurements; the combined visual panel matched the four WSRLR and 16 texture measurements in the entorhinal–hippocampal panel. Analyses retained identical training and testing participants and used ADNI3-only tuning. Holm correction covered eight additional comparisons. These regions served as anatomical comparators and were not assumed to be unaffected by AD.

**Supplementary Table 1.** Brain regions included in the MRI analyses.

| No. | Region |
|---|---|
| 1 | Banks of the superior temporal sulcus |
| 2 | Caudal anterior cingulate cortex |
| 3 | Caudal middle frontal cortex |
| 4 | Cuneus |
| 5 | Entorhinal cortex |
| 6 | Fusiform gyrus |
| 7 | Inferior parietal cortex |
| 8 | Inferior temporal cortex |
| 9 | Isthmus of the cingulate cortex |
| 10 | Lateral occipital cortex |
| 11 | Lateral orbitofrontal cortex |
| 12 | Lingual gyrus |
| 13 | Medial orbitofrontal cortex |
| 14 | Middle temporal cortex |
| 15 | Parahippocampal cortex |
| 16 | Paracentral lobule |
| 17 | Pars opercularis |
| 18 | Pars orbitalis |
| 19 | Pars triangularis |
| 20 | Pericalcarine cortex |
| 21 | Postcentral gyrus |
| 22 | Posterior cingulate cortex |
| 23 | Precentral gyrus |
| 24 | Precuneus |
| 25 | Rostral anterior cingulate cortex |
| 26 | Rostral middle frontal cortex |
| 27 | Superior frontal cortex |
| 28 | Superior parietal cortex |
| 29 | Superior temporal cortex |
| 30 | Supramarginal gyrus |
| 31 | Frontal pole |
| 32 | Temporal pole |
| 33 | Transverse temporal cortex |
| 34 | Insula |
| 35 | Hippocampus |
| 36 | Amygdala |

Thirty-six regions per hemisphere were analyzed (34 cortical parcels plus hippocampus and amygdala), yielding 72 unilateral regions.

**Supplementary Table 2.** Contributions of WSRLR and texture to external cognitive prediction

**A. External prediction performance across four feature sets**

| Score | Model | Baseline $R^2$ | Baseline + WSRLR $R^2$ | Baseline + texture $R^2$ | Baseline + both $R^2$ |
|---|---|---|---|---|---|
| MoCA | Ridge | 0.1588 | 0.1864 | 0.2205 | 0.2371 |
| MoCA | Elastic net | 0.1681 | 0.1920 | 0.2308 | 0.2439 |
| MoCA | Gradient boosting | 0.1165 | 0.1478 | 0.2174 | 0.2203 |
| MoCA | TabPFN-v2 | 0.1631 | 0.1765 | 0.2021 | 0.2135 |
| MMSE | Ridge | 0.1964 | 0.2319 | 0.2542 | 0.2690 |
| MMSE | Elastic net | 0.2044 | 0.2376 | 0.2714 | 0.2842 |
| MMSE | Gradient boosting | 0.0567 | 0.1191 | 0.1984 | 0.2136 |
| MMSE | TabPFN-v2 | 0.0867 | 0.0924 | 0.1748 | 0.1834 |

**B. Incremental value of WSRLR**

| Score | Model | WSRLR added to baseline $\Delta R^2$ (95% CI) | Holm P | WSRLR added to texture $\Delta R^2$ (95% CI) | Holm P |
|---|---|---|---|---|---|
| MoCA | Ridge | +0.0277 (0.0170 to 0.0391) | **0.0016** | +0.0166 (0.0088 to 0.0250) | **0.0016** |
| MoCA | Elastic net | +0.0239 (0.0136 to 0.0351) | **0.0016** | +0.0131 (0.0053 to 0.0214) | **0.0165** |
| MoCA | Gradient boosting | +0.0313 (0.0035 to 0.0585) | 0.3070 | +0.0030 (-0.0150 to 0.0201) | 1.0000 |
| MoCA | TabPFN-v2 | +0.0133 (0.0032 to 0.0238) | 0.1800 | +0.0114 (-0.0006 to 0.0242) | 0.4084 |
| MMSE | Ridge | +0.0355 (0.0180 to 0.0558) | **0.0016** | +0.0148 (0.0034 to 0.0273) | 0.1800 |
| MMSE | Elastic net | +0.0332 (0.0145 to 0.0547) | **0.0050** | +0.0128 (0.0020 to 0.0256) | 0.2808 |
| MMSE | Gradient boosting | +0.0625 (0.0112 to 0.1136) | 0.2730 | +0.0152 (-0.0099 to 0.0383) | 0.7065 |
| MMSE | TabPFN-v2 | +0.0058 (-0.0197 to 0.0311) | 1.0000 | +0.0086 (-0.0020 to 0.0190) | 0.5185 |

MoCA: n=868 (ADNI4, 422; NACC, 380; OASIS-3, 66). MMSE: n=551 (ADNI4, 485; OASIS-3, 66). Identical participants were used across feature sets within each outcome. ADNI3 training samples were n=639 for MoCA and n=650 for MMSE. Baseline inputs comprised age, sex, education, log intracranial volume, scanner-manufacturer indicators, and 36 log-transformed regional volume-to-intracranial-volume ratios. Added features comprised four T1/FLAIR WSRLR measurements and/or 16 GLCM texture measurements from the entorhinal cortex and hippocampus. In panel B, WSRLR added to texture compares baseline + both with baseline + texture. Abbreviations: ADNI, Alzheimer's Disease Neuroimaging Initiative; CI, confidence interval; FLAIR, fluid-attenuated inversion recovery; GLCM, gray-level co-occurrence matrix; MMSE, Mini-Mental State Examination; MoCA, Montreal Cognitive Assessment; NACC, National Alzheimer's Coordinating Center; OASIS, Open Access Series of Imaging Studies; WSRLR, within-scan regional-to-rest log ratio.

**Supplementary Table 3.** Cohort-specific external prediction of cognitive performance

**A. Montreal Cognitive Assessment**

| Test cohort | Model | n | Baseline $R^2$ | Expanded $R^2$ | $\Delta R^2$ (95% CI) | Baseline MAE | Expanded MAE | MAE reduction (95% CI) |
|---|---|---|---|---|---|---|---|---|
| ADNI4 | Ridge | 422 | 0.217 | 0.280 | 0.063 (0.022 to 0.107) | 2.880 | 2.733 | 0.147 (0.058 to 0.237) |
| ADNI4 | Elastic net | 422 | 0.224 | 0.287 | 0.063 (0.024 to 0.107) | 2.872 | 2.721 | 0.152 (0.069 to 0.236) |
| ADNI4 | Histogram gradient boosting | 422 | 0.190 | 0.275 | 0.085 (0.032 to 0.139) | 2.855 | 2.763 | 0.092 (−0.010 to 0.198) |
| ADNI4 | TabPFN v2 | 422 | 0.189 | 0.234 | 0.044 (−0.002 to 0.089) | 2.837 | 2.818 | 0.019 (−0.067 to 0.106) |
| NACC | Ridge | 380 | 0.014 | 0.124 | 0.109 (0.049 to 0.179) | 2.828 | 2.676 | 0.152 (0.055 to 0.250) |
| NACC | Elastic net | 380 | 0.029 | 0.130 | 0.101 (0.042 to 0.168) | 2.792 | 2.660 | 0.133 (0.039 to 0.226) |
| NACC | Histogram gradient boosting | 380 | −0.040 | 0.099 | 0.140 (0.050 to 0.232) | 2.854 | 2.683 | 0.171 (0.053 to 0.294) |
| NACC | TabPFN v2 | 380 | 0.066 | 0.124 | 0.059 (0.000 to 0.124) | 2.683 | 2.590 | 0.094 (−0.004 to 0.195) |
| OASIS-3 | Ridge | 66 | 0.082 | 0.152 | 0.070 (−0.046 to 0.291) | 2.040 | 2.021 | 0.019 (−0.137 to 0.183) |
| OASIS-3 | Elastic net | 66 | 0.090 | 0.171 | 0.081 (−0.041 to 0.315) | 2.041 | 1.999 | 0.042 (−0.121 to 0.208) |
| OASIS-3 | Histogram gradient boosting | 66 | −0.114 | 0.006 | 0.120 (−0.115 to 0.530) | 2.392 | 2.310 | 0.082 (−0.204 to 0.359) |
| OASIS-3 | TabPFN v2 | 66 | 0.127 | 0.226 | 0.099 (−0.033 to 0.355) | 2.161 | 2.015 | 0.147 (−0.018 to 0.319) |

**B. Mini-mental state examination**

| Test cohort | Model | n | Baseline $R^2$ | Expanded $R^2$ | $\Delta R^2$ (95% CI) | Baseline MAE | Expanded MAE | MAE reduction (95% CI) |
|---|---|---|---|---|---|---|---|---|
| ADNI4 | Ridge | 485 | 0.187 | 0.265 | 0.079 (0.020 to 0.143) | 1.656 | 1.551 | 0.105 (0.040 to 0.172) |
| ADNI4 | Elastic net | 485 | 0.197 | 0.282 | 0.085 (0.028 to 0.147) | 1.648 | 1.547 | 0.101 (0.038 to 0.166) |
| ADNI4 | Histogram gradient boosting | 485 | 0.060 | 0.227 | 0.167 (0.063 to 0.282) | 1.688 | 1.558 | 0.129 (0.050 to 0.216) |
| ADNI4 | TabPFN v2 | 485 | 0.052 | 0.163 | 0.111 (0.042 to 0.178) | 1.579 | 1.518 | 0.060 (0.004 to 0.121) |
| OASIS-3 | Ridge | 66 | 0.037 | 0.048 | 0.011 (−0.131 to 0.576) | 1.215 | 1.280 | −0.065 (−0.208 to 0.073) |
| OASIS-3 | Elastic net | 66 | 0.008 | 0.046 | 0.038 (−0.115 to 0.785) | 1.235 | 1.276 | −0.041 (−0.191 to 0.102) |
| OASIS-3 | Histogram gradient boosting | 66 | −0.357 | −0.287 | 0.070 (−0.402 to 1.437) | 1.491 | 1.453 | 0.038 (−0.189 to 0.241) |
| OASIS-3 | TabPFN v2 | 66 | 0.253 | 0.175 | −0.079 (−0.147 to 0.039) | 1.025 | 1.069 | −0.044 (−0.097 to 0.003) |

Baseline models were compared with models adding entorhinal and hippocampal WSRLR and texture. Positive $\Delta R^2$ and mean absolute error (MAE) reduction favor expanded models. Nominal 95% confidence intervals used 5,000 paired bootstrap resamples without adjustment for multiple comparisons. Abbreviations: ADNI, Alzheimer's Disease Neuroimaging Initiative; CI, confidence interval; FLAIR, fluid-attenuated inversion recovery; GLCM, gray-level co-occurrence matrix; MAE, mean absolute error; MMSE, Mini-Mental State Examination; MoCA, Montreal Cognitive Assessment; NACC, National Alzheimer's Coordinating Center; OASIS-3, Open Access Series of Imaging Studies, third release; $R^2$, coefficient of determination; WSRLR, within-scan regional-to-rest log ratio.

**Supplementary Table 4.** Calibration of external cognitive predictions

### A. Montreal Cognitive Assessment

| Test | Model | Predictor | n | Intercept | Slope | Mean error |
|---|---|---|---|---|---|---|
| ADNI4 | Ridge | Baseline | 422 | 3.87 (−0.13 to 7.82) | 0.813 (0.661 to 0.968) | 0.82 (0.47 to 1.18) |
| | | Expanded | | 1.25 (−2.96 to 5.30) | 0.915 (0.759 to 1.078) | 0.89 (0.55 to 1.23) |
| ADNI4 | Elastic net | Baseline | 422 | 3.55 (−0.47 to 7.49) | 0.826 (0.674 to 0.980) | 0.82 (0.47 to 1.18) |
| | | Expanded | | 0.77 (−3.55 to 4.90) | 0.934 (0.777 to 1.103) | 0.88 (0.54 to 1.22) |
| ADNI4 | Gradient boosting | Baseline | 422 | 5.14 (1.10 to 9.08) | 0.767 (0.615 to 0.925) | 0.67 (0.30 to 1.04) |
| | | Expanded | | 1.20 (−2.81 to 5.14) | 0.919 (0.766 to 1.072) | 0.85 (0.51 to 1.19) |
| ADNI4 | TabPFN-v2 | Baseline | 422 | 3.28 (−0.73 to 7.12) | 0.825 (0.679 to 0.978) | 1.17 (0.82 to 1.53) |
| | | Expanded | | 0.65 (−3.32 to 4.58) | 0.916 (0.770 to 1.068) | 1.50 (1.17 to 1.84) |
| NACC | Ridge | Baseline | 380 | 12.00 (7.47 to 16.48) | 0.533 (0.359 to 0.709) | −0.45 (−0.80 to −0.09) |
| | | Expanded | | 9.17 (4.40 to 13.35) | 0.644 (0.481 to 0.830) | −0.33 (−0.67 to 0.01) |
| NACC | Elastic net | Baseline | 380 | 11.61 (7.02 to 16.15) | 0.548 (0.371 to 0.725) | −0.42 (−0.77 to −0.06) |
| | | Expanded | | 8.82 (3.98 to 13.10) | 0.659 (0.492 to 0.847) | −0.35 (−0.68 to −0.01) |
| NACC | Gradient boosting | Baseline | 380 | 13.59 (9.30 to 17.62) | 0.467 (0.311 to 0.632) | −0.35 (−0.72 to 0.02) |
| | | Expanded | | 9.92 (4.77 to 14.76) | 0.610 (0.421 to 0.811) | −0.16 (−0.51 to 0.18) |
| NACC | TabPFN-v2 | Baseline | 380 | 10.53 (5.44 to 15.27) | 0.579 (0.397 to 0.774) | 0.13 (−0.22 to 0.48) |
| | | Expanded | | 8.81 (4.12 to 13.25) | 0.640 (0.473 to 0.818) | 0.40 (0.05 to 0.73) |
| OASIS-3 | Ridge | Baseline | 66 | 10.14 (1.43 to 20.08) | 0.639 (0.269 to 0.968) | −0.89 (−1.52 to −0.31) |
| | | Expanded | | 6.84 (−2.55 to 17.12) | 0.771 (0.375 to 1.129) | −1.00 (−1.57 to −0.45) |
| OASIS-3 | Elastic net | Baseline | 66 | 10.05 (1.33 to 20.12) | 0.642 (0.266 to 0.974) | −0.86 (−1.48 to −0.30) |
| | | Expanded | | 6.32 (−3.22 to 16.39) | 0.791 (0.406 to 1.156) | −0.98 (−1.54 to −0.44) |
| OASIS-3 | Gradient boosting | Baseline | 66 | 12.87 (2.72 to 23.26) | 0.537 (0.141 to 0.925) | −1.08 (−1.75 to −0.46) |
| | | Expanded | | 10.80 (−0.20 to 20.79) | 0.615 (0.234 to 1.039) | −0.93 (−1.56 to −0.33) |
| OASIS-3 | TabPFN-v2 | Baseline | 66 | 8.03 (−2.43 to 22.39) | 0.717 (0.172 to 1.113) | −0.71 (−1.32 to −0.13) |
| | | Expanded | | 6.17 (−5.30 to 18.22) | 0.777 (0.325 to 1.204) | −0.34 (−0.94 to 0.22) |

**B. Mini-Mental State Examination**

| Test | Model | Predictor | n | Intercept | Slope | Mean error |
|---|---|---|---|---|---|---|
| ADNI4 | Ridge | Baseline | 485 | 8.61 (4.56 to 12.70) | 0.688 (0.546 to 0.829) | 0.16 (−0.04 to 0.36) |
| | | Expanded | | 6.66 (2.81 to 10.13) | 0.755 (0.635 to 0.890) | 0.23 (0.04 to 0.42) |
| ADNI4 | Elastic net | Baseline | 485 | 8.22 (4.17 to 12.26) | 0.702 (0.562 to 0.845) | 0.15 (−0.05 to 0.35) |
| | | Expanded | | 5.97 (1.91 to 9.51) | 0.781 (0.659 to 0.923) | 0.18 (0.00 to 0.38) |
| ADNI4 | Gradient boosting | Baseline | 485 | 12.32 (8.13 to 16.34) | 0.557 (0.415 to 0.703) | 0.13 (−0.08 to 0.35) |
| | | Expanded | | 6.85 (1.40 to 11.56) | 0.748 (0.584 to 0.937) | 0.26 (0.07 to 0.46) |
| ADNI4 | TabPFN-v2 | Baseline | 485 | 8.62 (0.92 to 13.81) | 0.674 (0.496 to 0.943) | 0.70 (0.50 to 0.92) |
| | | Expanded | | 5.88 (−1.74 to 11.30) | 0.772 (0.585 to 1.035) | 0.65 (0.46 to 0.85) |
| OASIS-3 | Ridge | Baseline | 66 | 11.12 (−6.11 to 25.65) | 0.631 (0.130 to 1.228) | −0.62 (−0.98 to −0.22) |
| | | Expanded | | 7.91 (−13.04 to 24.43) | 0.747 (0.174 to 1.477) | −0.71 (−1.06 to −0.32) |
| OASIS-3 | Elastic net | Baseline | 66 | 11.39 (−5.62 to 25.60) | 0.623 (0.133 to 1.207) | −0.67 (−1.04 to −0.26) |
| | | Expanded | | 8.15 (−13.75 to 24.56) | 0.737 (0.169 to 1.495) | −0.68 (−1.03 to −0.27) |
| OASIS-3 | Gradient boosting | Baseline | 66 | 20.74 (0.78 to 34.22) | 0.294 (−0.172 to 0.989) | −0.71 (−1.15 to −0.20) |
| | | Expanded | | 19.41 (−2.72 to 29.75) | 0.341 (−0.013 to 1.104) | −0.68 (−1.10 to −0.19) |
| OASIS-3 | TabPFN-v2 | Baseline | 66 | −31.08 (−67.52 to 21.39) | 2.084 (0.272 to 3.340) | −0.23 (−0.55 to 0.18) |
| | | Expanded | | −8.89 (−75.53 to 23.90) | 1.316 (0.190 to 3.610) | −0.24 (−0.58 to 0.19) |

Baseline included demographic and volumetric inputs; expanded models additionally included entorhinal and hippocampal T1/FLAIR signal and texture. Calibration regressed observed on predicted scores; ideal intercept = 0 and slope = 1. Mean error = predicted minus observed score, in points; positive values indicate overestimation. Intervals are nominal 95% percentile CIs from 5,000 within-cohort participant bootstrap resamples, conditional on the fitted models. Predictions were not recalibrated.

Abbreviations: ADNI4, Alzheimer's Disease Neuroimaging Initiative phase 4; CI, confidence interval; FLAIR, fluid-attenuated inversion recovery; MMSE, Mini-Mental State Examination; MoCA, Montreal Cognitive Assessment; NACC, National Alzheimer's Coordinating Center; OASIS-3, Open Access Series of Imaging Studies, third release; TabPFN, Tabular Prior-data Fitted Network.

**Supplementary Table 5.** Discrimination of low cognitive scores after adding MRI signal and texture

**A. Montreal Cognitive Assessment <26**

| Test cohorts | Model | Predictor panel | Low / total n | AUROC (95% CI) | ΔAUROC (95% CI) | Holm-adjusted P |
|---|---|---|---|---|---|---|
| ADNI4, NACC, OASIS-3 | Ridge | Baseline | 421/868 | 0.654 (0.617 to 0.691) | 0.029 (0.017 to 0.043) | **<0.001** |
| | | Expanded | | 0.684 (0.647 to 0.718) | | |
| ADNI4, NACC, OASIS-3 | Elastic net | Baseline | 421/868 | 0.657 (0.620 to 0.693) | 0.029 (0.017 to 0.042) | **<0.001** |
| | | Expanded | | 0.686 (0.650 to 0.721) | | |
| ADNI4, NACC, OASIS-3 | Gradient boosting | Baseline | 421/868 | 0.646 (0.609 to 0.682) | 0.007 (−0.011 to 0.026) | 0.449 |
| | | Expanded | | 0.653 (0.616 to 0.689) | | |
| ADNI4, NACC, OASIS-3 | TabPFN-v2 | Baseline | 421/868 | 0.647 (0.608 to 0.683) | 0.026 (0.013 to 0.040) | **0.003** |
| | | Expanded | | 0.672 (0.636 to 0.708) | | |

**B. Mini-Mental State Examination <24**

| Test cohorts | Model | Predictor panel | Low / total n | AUROC (95% CI) | ΔAUROC (95% CI) | Holm-adjusted P |
|---|---|---|---|---|---|---|
| ADNI4, OASIS-3 | Ridge | Baseline | 35/551 | 0.877 (0.818 to 0.928) | 0.020 (−0.009 to 0.048) | 0.160 |
| | | Expanded | | 0.897 (0.841 to 0.945) | | |
| ADNI4, OASIS-3 | Elastic net | Baseline | 35/551 | 0.882 (0.825 to 0.930) | 0.028 (−0.001 to 0.056) | 0.091 |
| | | Expanded | | 0.909 (0.861 to 0.951) | | |
| ADNI4, OASIS-3 | Gradient boosting | Baseline | 35/551 | 0.793 (0.707 to 0.867) | 0.093 (0.042 to 0.148) | **<0.001** |
| | | Expanded | | 0.885 (0.814 to 0.941) | | |
| ADNI4, OASIS-3 | TabPFN-v2 | Baseline | 35/551 | 0.872 (0.801 to 0.925) | 0.036 (0.014 to 0.060) | **0.005** |
| | | Expanded | | 0.908 (0.842 to 0.956) | | |

Baseline: demographic and volumetric inputs. Expanded: baseline plus entorhinal and hippocampal T1/FLAIR WSRLR and GLCM texture. AUROC used negative frozen score predictions; no refitting or recalibration. Positive ΔAUROC favors expanded models. Nominal 95% CIs used 5,000 cohort-stratified paired bootstraps; P values used 20,000 paired prediction swaps with Holm correction across four models within each outcome.

Counts show low-score/total participants. MoCA cohorts: ADNI4 (422), NACC (380), OASIS-3 (66); MMSE: ADNI4 (485), OASIS-3 (66). Of 35 MMSE <24 cases, 34 were ADNI4 and one OASIS-3, limiting independent replication.

Abbreviations: ADNI4, Alzheimer’s Disease Neuroimaging Initiative phase 4; AUROC, area under the receiver operating characteristic curve; CI, confidence interval; FLAIR, fluid-attenuated inversion recovery; GLCM, gray-level co-occurrence matrix; MMSE, Mini-Mental State Examination; MoCA, Montreal Cognitive Assessment; NACC, National Alzheimer’s Coordinating Center; OASIS-3, Open Access Series of Imaging Studies; TabPFN, Tabular Prior-data Fitted Network; WSRLR, within-scan regional-to-rest log ratio.

### T1

| Region | ADNI3 | ADNI4 | MCSA | SCAN | CLARiTI | OASIS-3 |
|---|---|---|---|---|---|---|
| Banks of superior temporal sulcus | 0.55 | 0.76 | 0.92 | 0.66 | 0.83 | 0.74 |
| Caudal anterior cingulate | 0.61 | 0.64 | 0.94 | 0.55 | 0.60 | 0.89 |
| Caudal middle frontal | 0.86 | 0.78 | 0.90 | 0.69 | 0.80 | 0.93 |
| Cuneus | 0.70 | 0.83 | 0.87 | 0.76 | 0.76 | 0.86 |
| **Entorhinal** | 0.74 | 0.85 | 0.83 | 0.83 | 0.83 | 0.95 |
| Fusiform | 0.77 | 0.82 | 0.84 | 0.73 | 0.86 | 0.89 |
| Inferior parietal | 0.68 | 0.77 | 0.84 | 0.70 | 0.86 | 0.90 |
| Inferior temporal | 0.52 | 0.69 | 0.90 | 0.53 | 0.73 | 0.91 |
| Isthmus cingulate | 0.67 | 0.75 | 0.86 | 0.64 | 0.63 | 0.85 |
| Lateral occipital | 0.76 | 0.80 | 0.89 | 0.76 | 0.75 | 0.91 |
| Lateral orbitofrontal | 0.69 | 0.79 | 0.94 | 0.80 | 0.86 | 0.93 |
| Lingual | 0.74 | 0.82 | 0.85 | 0.67 | 0.76 | 0.84 |
| Medial orbitofrontal | 0.63 | 0.73 | 0.93 | 0.67 | 0.79 | 0.90 |
| Middle temporal | 0.64 | 0.63 | 0.90 | 0.51 | 0.70 | 0.87 |
| Parahippocampal | 0.76 | 0.77 | 0.76 | 0.72 | 0.77 | 0.95 |
| Paracentral | 0.70 | 0.78 | 0.96 | 0.81 | 0.82 | 0.92 |
| Pars opercularis | 0.69 | 0.79 | 0.84 | 0.51 | 0.79 | 0.77 |
| Pars orbitalis | 0.57 | 0.78 | 0.88 | 0.71 | 0.60 | 0.90 |
| Pars triangularis | 0.69 | 0.80 | 0.89 | 0.63 | 0.85 | 0.88 |
| Pericalcarine | 0.73 | 0.81 | 0.74 | 0.83 | 0.85 | 0.79 |
| Postcentral | 0.78 | 0.77 | 0.91 | 0.69 | 0.73 | 0.92 |
| Posterior cingulate | 0.59 | 0.73 | 0.97 | 0.67 | 0.68 | 0.83 |
| Precentral | 0.86 | 0.86 | 0.95 | 0.75 | 0.88 | 0.96 |
| Precuneus | 0.60 | 0.76 | 0.91 | 0.68 | 0.83 | 0.89 |
| Rostral anterior cingulate | 0.56 | 0.65 | 0.94 | 0.48 | 0.70 | 0.92 |
| Rostral middle frontal | 0.58 | 0.69 | 0.84 | 0.56 | 0.70 | 0.89 |
| Superior frontal | 0.70 | 0.65 | 0.87 | 0.62 | 0.67 | 0.90 |
| Superior parietal | 0.75 | 0.86 | 0.94 | 0.86 | 0.91 | 0.91 |
| Superior temporal | 0.62 | 0.67 | 0.88 | 0.65 | 0.74 | 0.85 |
| Supramarginal | 0.71 | 0.74 | 0.92 | 0.59 | 0.77 | 0.83 |
| Frontal pole | 0.48 | 0.77 | 0.86 | 0.74 | 0.71 | 0.88 |
| Temporal pole | 0.70 | 0.85 | 0.85 | 0.83 | 0.79 | 0.95 |
| Transverse temporal | 0.59 | 0.71 | 0.82 | 0.69 | 0.47 | 0.88 |
| Insula | 0.66 | 0.81 | 0.86 | 0.71 | 0.70 | 0.85 |
| Hippocampus | 0.65 | 0.73 | 0.73 | 0.64 | 0.64 | 0.86 |
| Amygdala | 0.65 | 0.76 | 0.76 | 0.72 | 0.71 | 0.87 |

#### FLAIR

| Region | ADNI3 | ADNI4 | MCSA | SCAN | CLARiTI | OASIS-3 |
|---|---|---|---|---|---|---|
| Banks of superior temporal sulcus | 0.70 | 0.78 | 0.88 | 0.73 | 0.79 | 0.69 |
| Caudal anterior cingulate | 0.53 | 0.67 | 0.92 | 0.51 | 0.78 | 0.82 |
| Caudal middle frontal | 0.55 | 0.65 | 0.86 | 0.67 | 0.67 | 0.86 |
| Cuneus | 0.55 | 0.65 | 0.77 | 0.46 | 0.75 | 0.88 |
| **Entorhinal** | 0.61 | 0.78 | 0.84 | 0.65 | 0.82 | 0.85 |
| Fusiform | 0.48 | 0.81 | 0.77 | 0.50 | 0.67 | 0.93 |
| Inferior parietal | 0.66 | 0.77 | 0.85 | 0.83 | 0.78 | 0.84 |
| Inferior temporal | 0.55 | 0.68 | 0.84 | 0.61 | 0.81 | 0.90 |
| Isthmus cingulate | 0.43 | 0.82 | 0.94 | 0.52 | 0.60 | 0.84 |
| Lateral occipital | 0.63 | 0.71 | 0.92 | 0.72 | 0.72 | 0.85 |
| Lateral orbitofrontal | 0.71 | 0.76 | 0.89 | 0.67 | 0.80 | 0.81 |
| Lingual | 0.43 | 0.61 | 0.64 | 0.38 | 0.62 | 0.82 |
| Medial orbitofrontal | 0.39 | 0.83 | 0.76 | 0.55 | 0.56 | 0.84 |
| Middle temporal | 0.45 | 0.60 | 0.86 | 0.43 | 0.67 | 0.80 |
| Parahippocampal | 0.43 | 0.64 | 0.81 | 0.48 | 0.58 | 0.79 |
| Paracentral | 0.52 | 0.65 | 0.86 | 0.45 | 0.53 | 0.80 |
| Pars opercularis | 0.73 | 0.72 | 0.85 | 0.70 | 0.74 | 0.78 |
| Pars orbitalis | 0.45 | 0.87 | 0.93 | 0.67 | 0.77 | 0.74 |
| Pars triangularis | 0.50 | 0.82 | 0.88 | 0.66 | 0.75 | 0.78 |
| Pericalcarine | 0.55 | 0.66 | 0.64 | 0.48 | 0.80 | 0.87 |
| Postcentral | 0.48 | 0.74 | 0.80 | 0.54 | 0.46 | 0.84 |
| Posterior cingulate | 0.46 | 0.77 | 0.95 | 0.49 | 0.73 | 0.76 |
| Precentral | 0.43 | 0.66 | 0.81 | 0.55 | 0.44 | 0.78 |
| Precuneus | 0.46 | 0.83 | 0.91 | 0.40 | 0.70 | 0.89 |
| Rostral anterior cingulate | 0.51 | 0.67 | 0.88 | 0.33 | 0.60 | 0.75 |
| Rostral middle frontal | 0.41 | 0.63 | 0.87 | 0.66 | 0.68 | 0.82 |
| Superior frontal | 0.56 | 0.64 | 0.90 | 0.55 | 0.54 | 0.90 |
| Superior parietal | 0.62 | 0.77 | 0.90 | 0.57 | 0.73 | 0.91 |
| Superior temporal | 0.62 | 0.71 | 0.89 | 0.56 | 0.75 | 0.77 |
| Supramarginal | 0.68 | 0.75 | 0.82 | 0.70 | 0.64 | 0.76 |
| Frontal pole | 0.37 | 0.78 | 0.84 | 0.70 | 0.66 | 0.92 |
| Temporal pole | 0.70 | 0.81 | 0.86 | 0.72 | 0.84 | 0.85 |
| Transverse temporal | 0.38 | 0.73 | 0.74 | 0.42 | 0.68 | 0.90 |
| Insula | 0.26 | 0.44 | 0.52 | 0.33 | 0.39 | 0.71 |
| Hippocampus | 0.46 | 0.59 | 0.61 | 0.44 | 0.62 | 0.71 |
| Amygdala | 0.49 | 0.56 | 0.53 | 0.47 | 0.69 | 0.71 |

**Supplementary Figure 1.** Within-group Spearman correlations (ρ) between regional WSRLR and CN-reference normalization methods across different cohorts. The data represent all 36 bilateral brain regions, with each WSRLR target referenced to the other 35 regions. Comparisons used the same 2,314 complete paired datasets for T1-weighted and FLAIR sequences. Abbreviations: ADNI3/4, Alzheimer's Disease Neuroimaging Initiative, phases 3 and 4; ADRD, Alzheimer's disease and related dementias; CLARiTI, Consortium for Clarity in ADRD Research Through Imaging; CN, cognitively normal; FLAIR, fluid-attenuated inversion recovery; MCSA, Mayo Clinic Study of Aging; NACC, National Alzheimer's Coordinating Center; OASIS-3, Open Access Series of Imaging Studies, third release; SCAN, Standardized Centralized Alzheimer's and Related Dementias Neuroimaging; T1, longitudinal relaxation time; WSRLR, within-scan regional-to-rest log ratio; ρ, Spearman rank correlation coefficient.

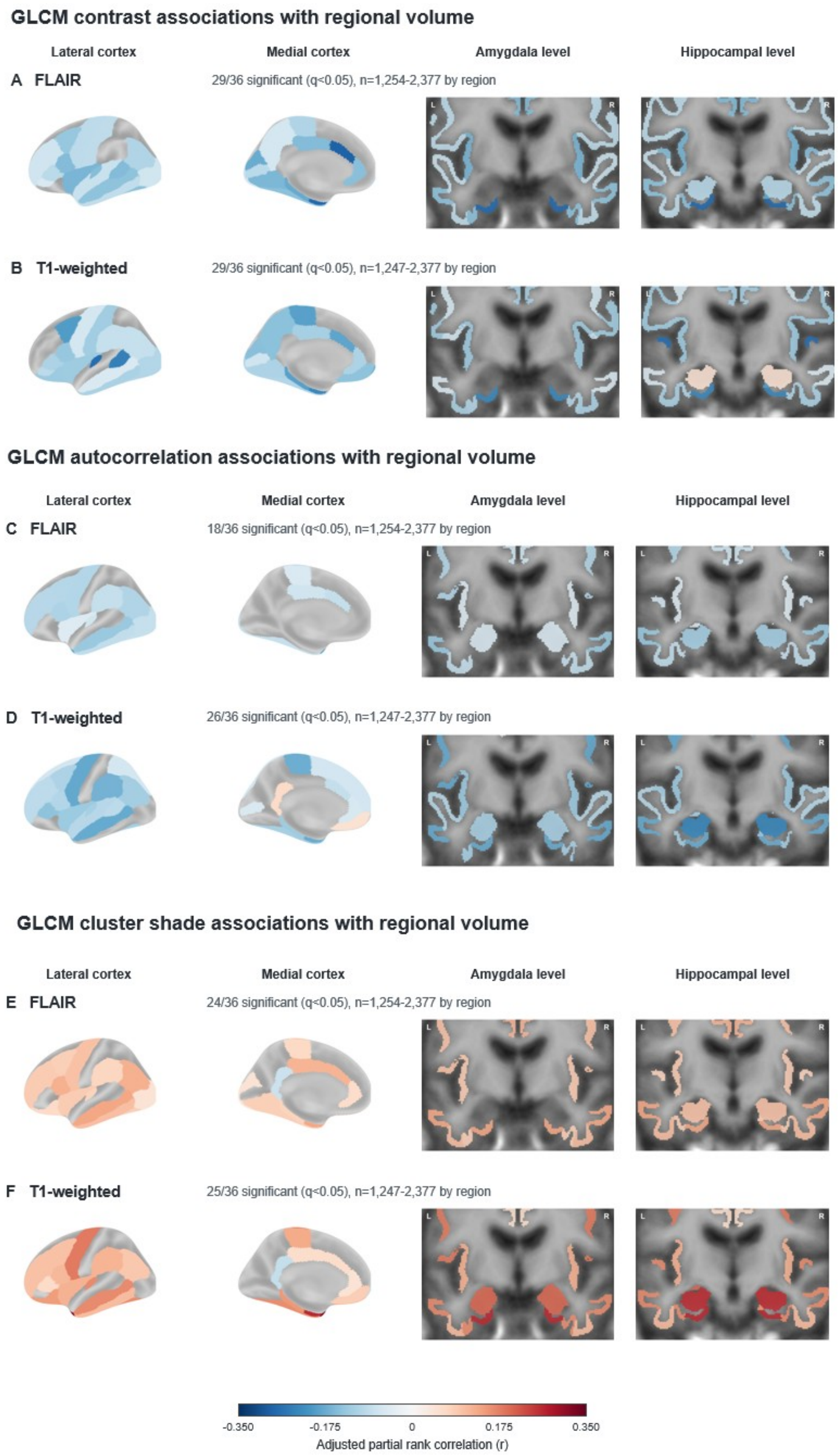


**Supplementary Figure 2.** Regional texture associations with corresponding brain volume. Pooled, cohort-stratified partial rank correlations are shown for GLCM contrast (A/B), autocorrelation (C/D), and cluster shade (E/F), with FLAIR/T1 paired. Models were adjusted for age, sex, education, ICV, and scanner; regional volumes were normalized to ICV. Colors indicate Benjamini-Hochberg FDR-adjusted q<0.05: red positive, blue negative. Abbreviations: FDR, false discovery rate; FLAIR, fluid-attenuated inversion recovery; GLCM, gray-level co-occurrence matrix; ICV, intracranial volume.

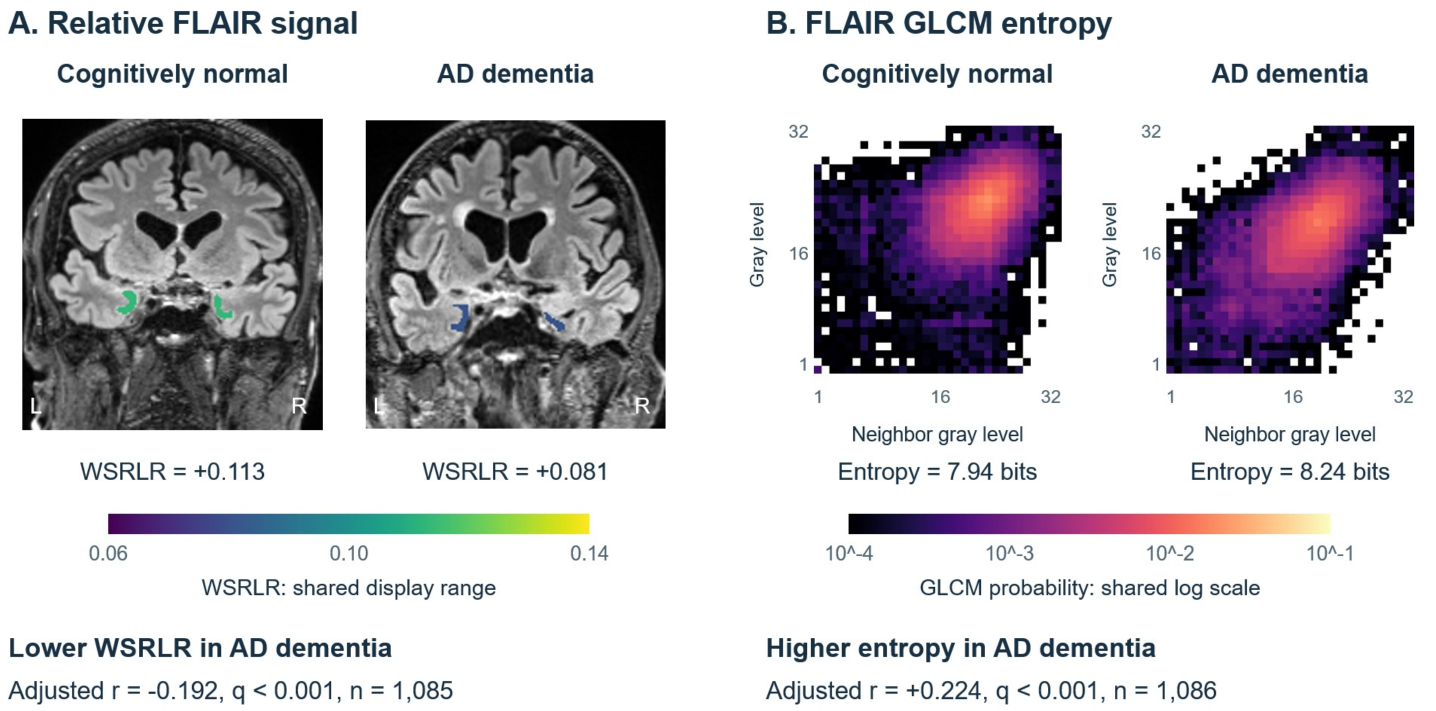


**Supplementary Figure 3.** Representative cases demonstrating entorhinal FLAIR signal and texture differences between cognitively normal (CN) and clinically attributed Alzheimer's disease dementia (AD). (A) Coronal FLAIR images show the relative signal in the entorhinal cortex within-scan regional-to-rest log ratio (WSRLR), which is relatively lower in the participant with AD compared to CN. WSRLR uses the other 35 bilateral gray-matter regions as the target-excluded reference. (B) Corresponding three-dimensional gray-level co-occurrence matrices (GLCMs) show increased entorhinal entropy in the AD participant, compared to CN. Statistics beneath each panel reflect pooled partial rank associations adjusted for age, sex, education, intracranial volume, corresponding regional volume, and scanner characteristics. Benjamini-Hochberg false discovery rate correction was used to calculate q-values. Abbreviations: FDR, false discovery rate; FLAIR, fluid-attenuated inversion recovery; WSRLR, within-scan regional-to-rest log ratio.

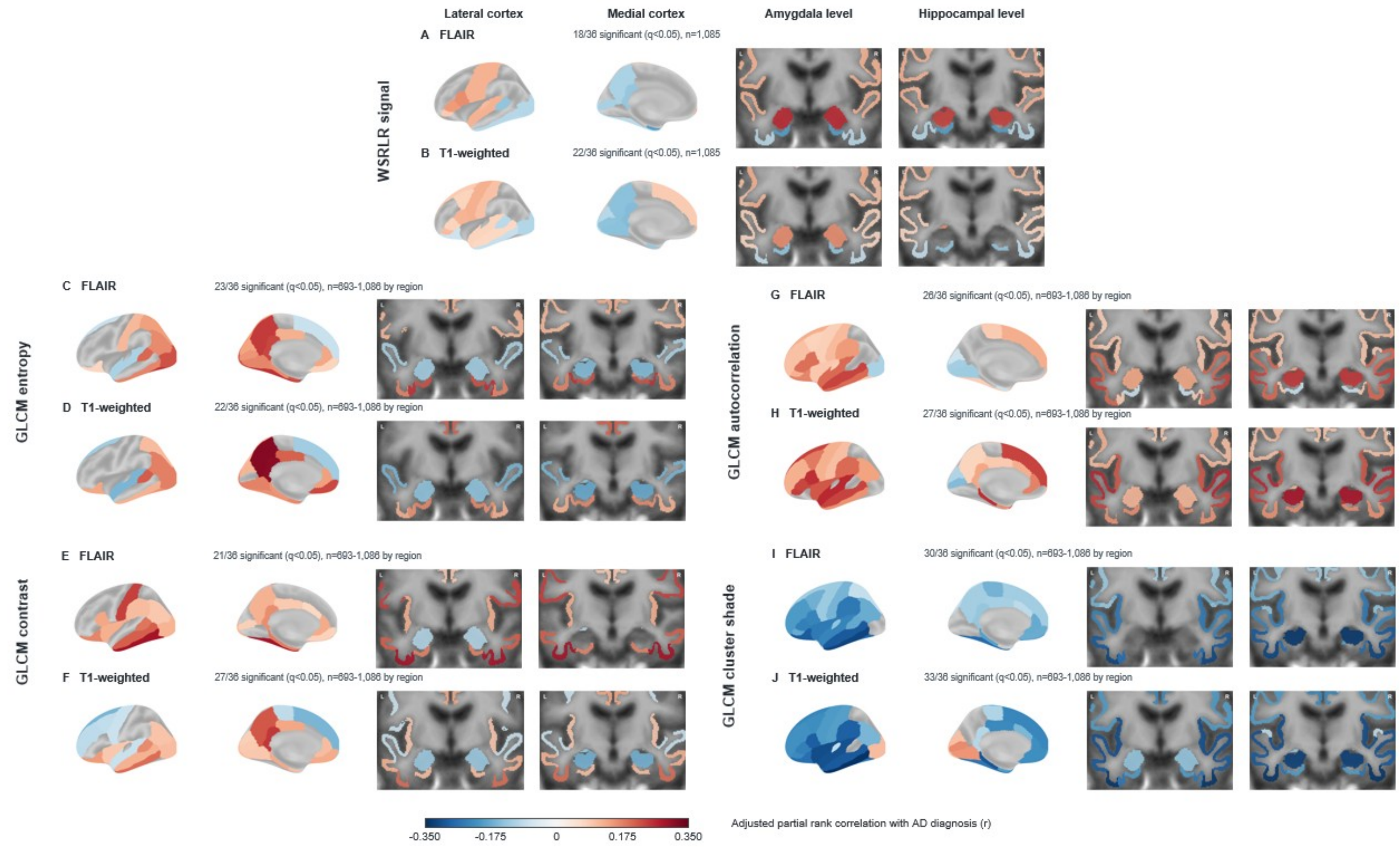


**Supplementary Figure 4.** Regional WSRLR signal and GLCM texture differences between clinically attributed Alzheimer's disease dementia and cognitively normal participants. Pooled, covariate-adjusted partial rank correlations are shown for WSRLR signal (A, B), GLCM entropy (C, D), contrast (E, F), autocorrelation (G, H), and cluster shade (I, J). Models included corresponding regional volume and other covariates. Colors encode adjusted partial rank association strength, with red indicating higher and blue lower measurements in AD dementia than in cognitively normal participants. WSRLR associations were significant in 18 FLAIR and 22 T1 regions. Only regions with Benjamini-Hochberg FDR-adjusted $q<0.05$ are colored. Abbreviations: FDR, false discovery rate; FLAIR, fluid-attenuated inversion recovery; GLCM, gray-level co-occurrence matrix; WSRLR, within-scan regional-to-rest log ratio.

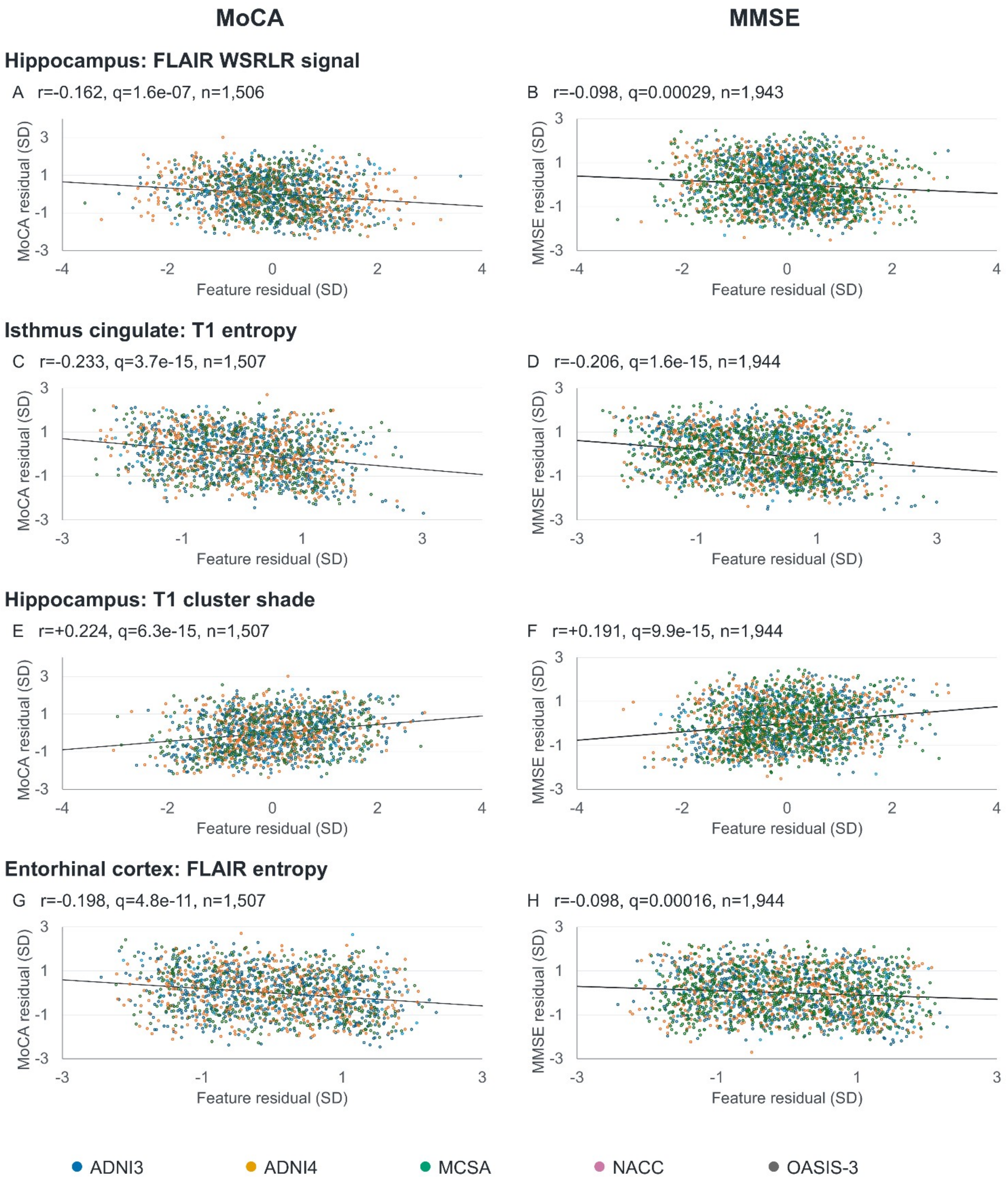


**Supplementary Figure 5.** Volume-adjusted regional signal and texture associations with cognition. Left/right: Montreal Cognitive Assessment (MoCA)/Mini-Mental State Examination (MMSE). Points show standardized rank residuals; colors identify cohorts and lines show pooled associations. Annotations report partial r, FDR-adjusted q, and n. MoCA analyses included ADNI3, ADNI4, NACC, and OASIS-3 (n=1,506–1,507); MMSE analyses included ADNI3, ADNI4, MCSA, and OASIS-3 (n=1,943–1,944).
